# Determinants and spatio-temporal structure of variability in wastewater SARS-CoV-2 viral load measurements in Switzerland: key insights for future surveillance efforts

**DOI:** 10.1101/2025.05.09.25327230

**Authors:** Julien Riou, Anna Fesser, Moritz Wagner, Katrin Schneider, Natalia Güdel-Krempaska, Christoph Ort, Tim Julian, Tanja Stadler, James D. Munday

**Author notes:** Corresponding authors: Current Address: Route de la Corniche 10, 1010 Lausanne, Switzerland & Schanzenstrasse 44, CH-4056 Basel, Switzerland Current &. These authors contributed equally to this work (Christoph Ort, Tim Julian, Tanja Stadler).

## Abstract

**Background:** Wastewater-based surveillance (WBS) has emerged as a valuable tool for monitoring the circulation of infectious pathogens in the population, offering a complement to classical surveillance methods. However, the interpretation of WBS data is often challenged by substantial variability and bias in viral load measurements, which can stem from differences in laboratory protocols, population demographics, and in-sewer fate and transport processes.

**Methods and Findings:** We analysed 23,025 wastewater samples collected between February 2022 and November 2023 from 118 wastewater treatment plants (WWTPs) across Switzerland. Samples of influent wastewater were processed by 8 independent laboratories using distinct concentration, extraction and quantification methods to estimate SARS-CoV-2 concentrations. Concentrations were converted to daily viral loads using wastewater flow rates, and normalized by the size of the population served by the treatment plant. To characterise the contributions of different sources of variation, we applied a Bayesian modelling framework, incorporating fixed effects and spatio–temporal random effects to separate the contributions of laboratory protocols, demographic factors, and geographic structure to the observed variability in SARS-CoV-2 viral loads. Our analysis revealed that laboratory-specific biases were substantial, and that local demographic characteristics (particularly the age structure of the catchment population) also influenced viral load estimates. Adjusting for these sources of bias improved the reliability of interpretations based on viral loads, as indicated by an increased correlation with regional COVID-19 hospitalization data (from 0.55 for raw data to 0.77 for adjusted temporal trends). Dynamic time warping clustering of the adjusted temporal trends uncovered distinct geographic clusters, highlighting persistent spatial structures that evolved over successive epidemic phases.

**Conclusions:** Our study demonstrates that variability in WBS data during the 2022-2023 Swiss national SARS-CoV-2 surveillance campaign is driven by a complex interplay of laboratory methods, population demographics, and spatio-temporal dynamics. Standardization of laboratory protocols and the implementation of robust statistical adjustment methods such as the one demonstrated here can enhance the reliability of WBS as a public health surveillance tool. These findings provide underscore the importance of advanced data processing methods for enhancing future surveillance efforts and for the effective integration of wastewater data into public health decision-making frameworks.

## 1 Introduction

Wastewater-based surveillance (WBS) is playing a an increasingly important role in many public health surveillance systems of infectious diseases [1]. While the analysis of wastewater concentrations of drugs, pharmaceuticals, and other biomarkers at wastewater treatment plants (WWTPs) has been used for decades, the application of this approach to produce real-time indicators of the prevalence of pathogens within communities is a fairly recent development, fuelled by the increased needs for surveillance during the SARS-CoV-2 pandemic [2]. As of 2023, WBS was being used to this aim in at least 43 countries [3]. This increase in popularity can be explained by several factors. Classical clinical surveillance programs are based on the notification of incident cases, hospitalisations and deaths with a positive laboratory test, the completeness and reliability of which can be influenced by age, sex/gender, and socioeconomic position. In contrast WBS is a population-based approach that does not depend on testing protocol or notifications [4–6]. Because it is exempt from these selection biases, the ambition of WBS programmes is to provide a more representative view of the population living in selected WWTP catchment areas, capturing the signal generated by infected people shedding sufficient virus, including people with no or mild symptoms. Considering its broad reach, WBS is also less costly than large-scale individual testing programmes. WBS can be applied to the general population by testing municipal sewage systems, or targeted at settings of particular importance such as hospitals [7], schools [8], university campuses [9] or airports [10]. Besides providing indirect information about epidemic dynamics, WBS has also been used to track the changing genomic landscapes of SARS-CoV-2, allowing population-level estimates of the relative abundance of variants circulating in the population [11]. Sequencing has also lead to earlier detection of variants than clinical sequencing [9, 12].

WBS relies upon the repeated collection, processing, and analysis of influent wastewater samples, generally in a fixed set of WWTPs. After collection, wastewater samples are processed to concentrate and extract nucleic acid and reduce co-concentrated impurities, and used as input to quantitative molecular detection methods, typically quantitative (qPCR) or digital (dPCR) polymerase chain reaction. The result is an estimate of the gene copies (gc) per unit volume of wastewater, which can then be scaled to daily viral loads using measurements of wastewater flow rates on each particular day (that can vary due many factors including rainfall and variable agricultural or industrial outflow). Other types of scaling can be applied to account for sources of variation that influence the relative proportion of human faecal inputs into the wastewater. These include total suspended solids or in-situ human faecal source markers such as Pepper Mild Mottle Virus (PMMoV) and CrAssphage RNA concentrations which consistently populate the human gut [13]. In addition, the scaled values are typically reported as population-normalised viral load. Normalisation allows for comparisons over time and space, which is key in the use of these data to track the dynamics of infection prevalence in the monitored population [14–16]. It also allows for the computation of reproduction numbers [17], commonly used to assess trends in this context. But a deeper look into the mechanisms of generation of wastewater data reveals many uncertainties in the path leading from pathogen transmission events in the population to wastewater viral load data. First, faecal shedding upon infection with SARS-CoV-2 dominates the inflow of viral RNA into community-level wastewater compared to other routes such as sputum, saliva or urine [18], and varies substantially between infected individuals and over the course of infection. The probability of faecal shedding is only about 40-50% on average, but is higher in case of gastrointestinal symptoms [19, 20], which can vary between variants [21]. Virus titre in faeces can vary widely according to individual characteristics such as age and comorbidities [22]. Second, the population that sheds into the catchment system of WWTP is not as clearly defined as its resident population, with movements during the day due to commuting and work, and periodic changes within weeks and years depending on events like holiday periods. A third source of variability may be variation within and between laboratory protocols (e.g., standard curves, N or S protein targets) and protocols (e.g., longer storage time that can lead to RNA degradation, sampling procedures) that can influence measured values [23–25]. Last, other environmental factors not covered by flow rate scaling may also play a role (e.g., changes in the chemical composition of the water). A direct consequence of these observations is that wastewater viral load data suffer from high noise and bias that vary between and within local contexts.

In this study, we take advantage of the dense network of WWTPs participating in SARS-CoV-2 surveillance in Switzerland between February 2022 and November 2023 to disentangle the different sources of variability and bias, and characterise the local determinants and the space-time structure of measured SARS-CoV-2 wastewater viral load. We then use these insights to produce adjusted temporal trends at the national and regional levels, corresponding to the underlying dynamics of SARS-CoV-2 infection after accounting for all other known sources of variability and bias. We then apply time-series clustering techniques to our de-biased estimates to evaluate the presence of geographic structures within the wastewater measurements from across Switzerland and support optimal monitoring site selection.

## 2 Methods

### 2.1 Viral concentration in wastewater

Data consist of wastewater samples collected between 7 February 2022 and 21 November 2023 in 118 WWTPs participating to the national WBS programme coordinated by the Swiss Federal Institute of Aquatic Science and Technology (Eawag), the Kantonal Laboratory of Zurich and the Swiss Federal Office of Public Health (FOPH). To facilitate the interpretation the study period was divided into five phases based on low points between epidemic waves (16 May 2022, 5 September 2022, 2 January 2023 and 3 July 2023). A total of 118 WWTPs, covering all 26 cantons, participated in the programme (Figure 1A), including six WWTPs directly handled by Eawag (with an internal laboratory, thereafter called *laboratory A*) that participated for the whole period (Table 1). WWTPs were identified by a unique number, by the canton and by the NUTS-2 region (7 higher-level administrative areas in Switzerland based on Eurostat’s *Nomenclature des Unités Territoriales Statistiques*). The minimal length of participation for a WWTP was 107 days. The mean frequency of sampling varied between 1.7 and 6.5 times per week depending on the WWTP (WWTPs with larger catchment populations were sampled more often). For each sample the day of collection was recorded, along with information about whether it fell on a Saturday or Sunday, or on a national holiday.

**Fig 1.**
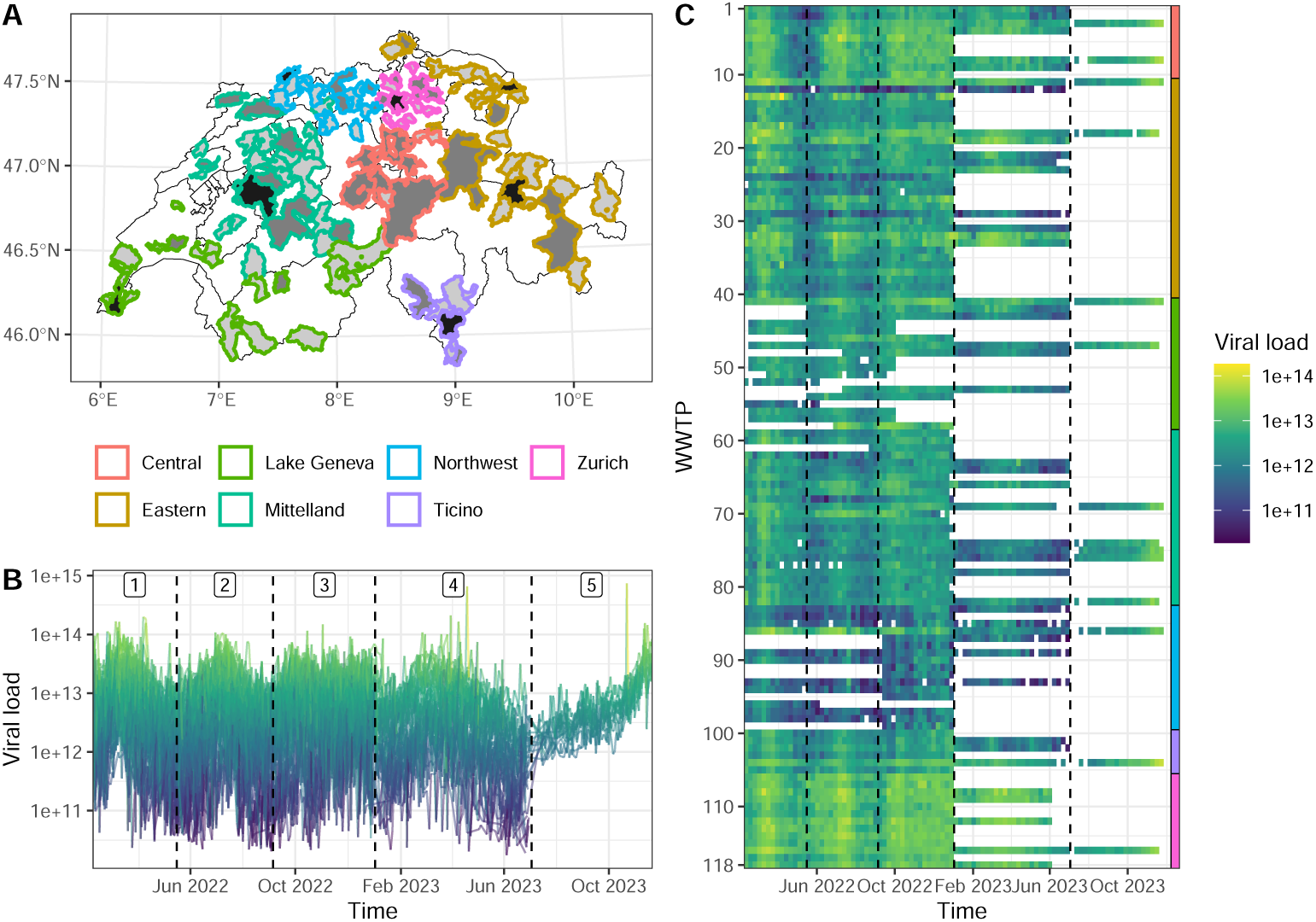
(A) Catchment areas of wastewater treatment plants [WWTPs] participating to SARS-CoV-2 surveillance by region, with the number of measurements in each WWTP over the study period (light grey *<*200, grey 200 to 500, black *≥*500). (B) All measurements of SARS-CoV-2 viral load in wastewater (values below the limit of quantification are removed; labels 1 to 5 correspond to *ad hoc* periods aligned with epidemic waves). (C) Weekly median SARS-CoV-2 viral load by WWTP (WWTPs are numbered alphabetically within regions indicated by the coloured scale on the right side; vertical dashed lines delimit the five periods labelled in panel B).

**Table 1.** Description of wastewater data used for SARS-CoV-2 surveillance in Switzerland (2022-2023).

|  | Value (count or median and range) |
| --- | --- |
| Number of WWTPs at peak | 118 |
| Number of laboratories | 8 |
| Number of laboratory methods | 10 |
| Total measurements | 23,025 |
| Measurements below LOQ | 675 (3%) |
| Measurements below LOD | 110 (0.5%) |
| Date of first measurement | 2022-02-07 |
| Date of last measurement | 2023-11-21 |
| Viral concentration [gc/L] | 94,350 (range: <LOD to 18,162,400) |
| Wastewater flow [m3/day] | 14,343 (range: 291 to 913,343) |
| Viral load [gc/day/100,000] | 3.7e+12 (range: <LOD to 7.4e+14) |
| Population covered | 37,954 (range: 2,052 to 479,727) |
| Proportion of population aged under 20 | 24% (range: 18% to 34%) |
| Proportion of population aged over 65 | 37% (range: 22% to 69%) |
| Median Swiss index of socio-economic position (SEP) | 63 (range: 54 to 78) |
| Employment factor | 0.51 (range: 0.23 to 1.12) |

Samples were stored at the wastewater treatment plant at 4*^o^*C and transported in ice-packed batches to one of 8 different laboratories (noted by letters A to H) for testing. Laboratories used different methods for concentration, extraction and quantification with qPCR/dPCR, and two laboratories reported a change in their protocol during the study period (noted by numbers, C1/C2 and G1/G2). This information was used to create a composite variable (laboratory *×* method, 10 levels) that was included in the statistical analysis. Measurements below the theoretical limit of quantification (LOQ) and the limit of detection (LOD), which represent limits of unacceptable quantification and detection uncertainty, respectively, were flagged in order to be treated differently in the analysis. The flow of wastewater on the day of the sampling was also reported by the WWTPs, and used to convert the measurements to daily viral load estimates. More details about the processing of wastewater samples can be found elsewhere [17, 26, 27].

### 2.2 Characteristics of WWTPs

We obtained the following datasets from the Swiss Federal Statistical Office (FSO) to produce variables characterising each WWTP. This was done by combining fine-level geographical information with the precise geographical shapes of WWTP catchment areas generated by Eawag and FOPH based on cantonal data, and consolidated by the Federal Office of the Environment. We used FSO’s *Population and Households Statistics* (STATPOP 2022) [28] by hectare to compute the total resident population (used to normalise viral loads, section 2.3), and the proportion of the population aged under 20 or over 65. We used FSO’s *Structural Business Statistics* (STATENT 2022) [29] to compute the number of full-time equivalent jobs per population within each WWTP (thereafter called *employment factor* ). To approximate the relative socio-economic position of each WWTP catchment, we used the Swiss index of socio-economic position (Swiss-SEP, based on FSO data [30] and updated in 2023) which is calculated for each building, and took the median value weighted by building occupancy based on the hectare-level population data. Last, we used the geographical shapes of WWTPs to compute an adjacency matrix, identifying WWTPs that have at least one shared border. Catchments with no contiguous borders were considered isolated from other included WWTPs. We used R with packages spdep and sf to manipulate geographical data [31].

### 2.3 Analysis of sources of variation in viral load

Measurements of viral concentration (*C*, unit: gene copies [gc] per liter) were converted into viral load (*V* ; unit: gc per day per 100,000 people) using the flow of wastewater on the same day (*F* ; unit: liters per day) and normalised by the size of the population of the WWTP catchment area (*P*):

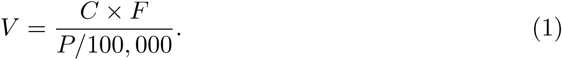

We use gamma regression to model viral load by WWTP over time, with a logarithmic link function that implies that all estimated effects are multiplicative. Model building was done incrementally, creating and comparing multiple versions based on domain knowledge, model fit and Watanabe-Akaike Information Criterion (WAIC) [32]. For the main analysis the mean of the selected model for WWTP *i*, region *j* and day *t* can be expressed as:

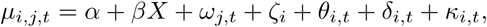

with intercept *α* and a vector of fixed effects *β* with covariate matrix *X* which included covariates for weekends (binary), national holidays (binary), laboratory method (10 levels, reference laboratory A, Eawag), proportion of the population aged under 20 (continuous) or over 65 (continuous), employment factor (continuous) and median Swiss-SEP (socio-economic position measure, continuous). All continuous covariates were centred and scaled by their standard deviation. The model also included a random walk of order 2 to model the underlying dynamics of viral load over time in each of the seven regions (*ω_j,t_*). This regional time trend was shifted up or down in each WWTP (*ζ_i_*) using a re-parameterised Besag-York-Mollié model (BYM2) [33]. This approach includes an Intrinsic Conditional Auto-Regressive (ICAR) component to account for the neighbourhood structure of WWTPs, allowing for higher correlation between shifts in neighbouring WWTPs than between non-adjacent areas, with a mixing parameter *ϕ* measuring the proportion of variance that can be attributed to this spatial structure (as opposed to an unstructured random effect). In addition to this WWTP-level shift, we used a random walk of order 1 to capture temporary deviations from the regional trend (*θ_i,t_*). We included additional variability for measurements below the LOD and below the LOQ using *i.i.d.* random effects (*δ_i,t_* and *κ_i,t_*).

We used Bayesian inference and integrated nested Laplace approximation as available in the R package INLA [34]. For prior distributions, we chose penalised complexity priors for the random walk and BYM2 components [35] and normal priors with mean 0 and variance 5 for fixed effects (corresponding to a 95% central range of 0.01 to 80 on the exponential scale). We report posterior estimates on the exponential scale as fold-change (FC) with 95% credible intervals. We use the fitted model to produce adjusted temporal trends in viral load using posterior estimates of *ω_j,t_*. We computed the Pearson correlation between *ω_j,t_* and weekly laboratory-confirmed SARS-CoV-2 hospitalisations in the same region, by period. We also report the posterior estimates of the adjusted bias in viral load by WWTP (*ζ_i_*). More details about the data analysis can be found in the Supplementary Text S1.

### 2.4 Clustering of WWTPs based on adjusted temporal trends in viral loads

To compare the variation in viral load between treatment plants, we adapted the model by removing the spatial and regional components:

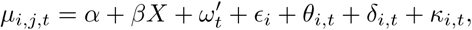

where *ω_t_^′^* is now a unique national temporal trend, and *ɛ_i_* is an *i.i.d.* random effect which allows temporally invariant deviation from the global trend. In effect, the local temporal component (*θ_i,t_*) measures the normalised deviation of viral loads from the global trend, when accounting for the other covariates in the model. We compared the posterior estimates for these variations for each phase of the study period by calculating the dynamic-time-warping (DTW) distance between the mean estimates, constructing a distance matrix between all WWTPs [36, 37]. We constructed a similarity matrix from the inverse DTW distances between time-series, applied the unweighted pair group method with arithmetic mean (UPGMA) hierarchical clustering algorithm and analysed the resultant cluster membership composition.

To assess the strength of connections within each cluster, we considered the average DTW distance between cluster members. Further, we characterised if a WWTP was closely connected to its cluster if the median DTW distance to members of their own cluster is smaller than the DTW distance to the closest five percent of WWTPs outside their cluster.

We assessed how similar communities were to each other by calculating the distance between the cluster centroids, with shorter distances between centroids indicating more similar communities.

## 3 Results

There was a total of 23,025 measurements of SARS-CoV-2 viral load in wastewater, including 110 (0.5%) below the LOD and 675 (3%) below the LOQ (Table 1). Measurements above the LOQ had high variability, ranging from 1.7e+10 to 7.4e+14 gc per day per 100,000 people, which corresponds to a ratio of 44,000 between the maximum and minimum values. There was also a lot of variability between WWTPs in the average viral load reported, with a ratio of 71 between the WWTPs with the highest and lowest mean load over the whole study period. Viral load also varied markedly over time, with the five successive waves highlighted in the methods section (Figure 1B). The number of participating WWTPs changed over time, with more than 100 WWTPs participating during phases 1 to 3 (maximum 115 during phase 3), downscaled to 46 in phase 4 and only 14 in phase 5. WWTPs situated in the same administrative area often displayed similar viral load dynamics over time on visual inspection, as can be clearly seen for numbers 1–10, 32–40, 71–83 or 106–118 (Figure 1C).

Taking advantage of the contrasts within the dense network of WWTPs, it was possible to estimate several fixed effects associated with viral load while adjusting for the variability over time and space. First, viral load measurements were heavily influenced by the laboratory and the methods used for the laboratory analyses (Figure 2A). For example, laboratory C1 reported measurements on average 33 times lower than the reference laboratory A (fold change [FC] 0.03, 95% credible interval [CrI] 0.02 to 0.05). This was only marginally improved by a change in laboratory methods occurring in October 2022 (laboratory C2, FC 0.09, 95%CrI 0.06 to 0.15). Laboratories E, F, G and H also reported on average far lower values than the reference laboratory, while only laboratories B and D were in agreement with the reference. Second, some characteristics of the population residing in the WWTP catchment area also influenced the measurements (Figure 2B). In particular, the age structure of the population appeared as an important driver, with a decrease of 7% in viral load for each standard deviation increase in the proportion of the population aged below 20 (FC 0.93, 95%CrI 0.80 to 1.04) and an increase of 11% for each standard deviation increase in the proportion aged above 65 (FC 1.11, 95%CrI 0.97 to 1.28). There were also modest increases with the number of full-time equivalent jobs per population within each WWTP – the so-called employment factor – or the median Swiss-SEP index were associated with changes in viral load (FC 1.09, 95%CrI 0.97 to 1.21 and 1.06, 95%CrI 0.93 to 1.21, respectively). Third, there was no signal indicating a systematic change in viral load during week-ends or during public holidays (Figure 2C).

**Fig 2.**
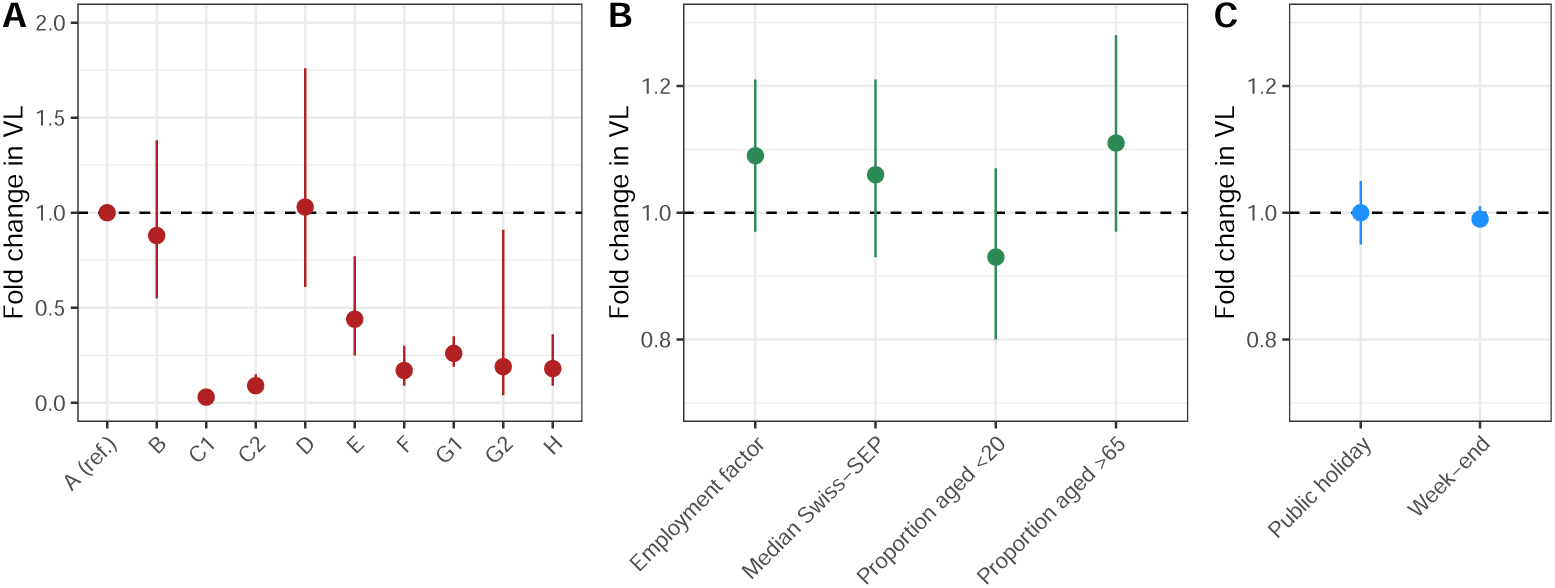
(A) Association between laboratories or changes in methods within laboratories and SARS-CoV-2 wastewater viral load [VL] (reference is laboratory A). (B) Association between characteristics of the population living in the wastewater treatment plants catchment area and VL (per unit-increase in standard deviation). (C) Association between calendar changes and VL (reference is no public holiday and week day, respectively).

The inferred space-time components also brought interesting insights. After adjusting for the fixed effects described above, the estimated time series followed similar trajectories across all seven regions, with the exception of few noticeable differences (Figure 3A). Compared to other regions, there were higher peaks of viral load in the Northwest region during phases 1 and 2, while on the contrary viral load measurements were comparatively lower in the Central region. During phase 3 the concordance between regions was relatively high, and higher levels of heterogeneity reappeared during phase 4 (with the Zurich region being particularly high) and phase 5 (with higher viral loads in the Eastern and Mittelland regions). The correlation between these adjusted time trends and the incidence of laboratory-confirmed SARS-CoV-2 hospitalisations within the same regions was overall very high (median 0.77, Figure 3B) and notably higher than correlations when the unadjusted viral load estimates were used (median 0.55, lower in 91% of the cases, S1 Text). Correlation was especially high during phases 1 and 2 (median 0.82 and 0.88, respectively) with one exception (Lake Geneva region, 0.41), then lower during phase 3 (median 0.43), and increased again during phases 4 and 5 (median 0.74 and 0.89, respectively). For phase 5, during which all analyses where centralized at laboratory A (Eawag), the correlation between hospitalizations and raw viral load measurements was also higher (0.80).

**Fig 3.**
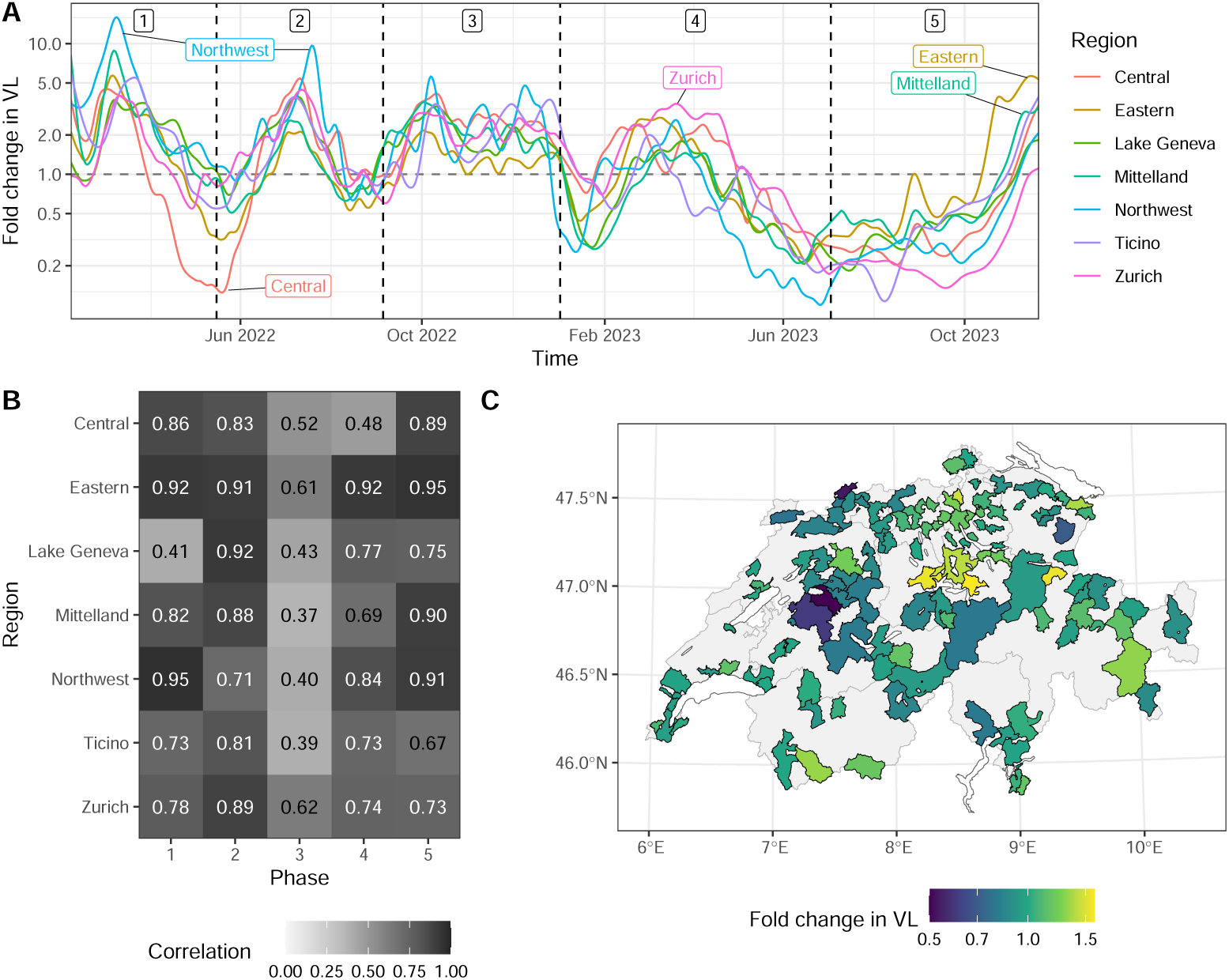
(A) Adjusted time trends in SARS-CoV-2 wastewater viral load in seven regions of Switzerland (uncertainty is not shown for clarity). (B) Correlation between adjusted time trends in viral load and laboratory-confirmed SARS-CoV-2 hospitalisations in the same region. (C) Adjusted bias in viral load by wastewater treatment plant.

Cross-correlation analyses indicated that the adjusted time trends were mostly synchronous or 1 week ahead of hospital admissions during periods 1, 2 and 3 (S1 Text). During periods 4 and 5, as COVID-19-related hospital admissions declined, the degree of synchronicity decreased. Last, we also observed some remaining bias at the level of the WWTP, even after adjusting for all the effects discussed above (Figure 3C). This remaining bias was relatively small, with a ratio of about 3 between the smallest fold change estimate of 0.50 (95%CrI 0.40 to 0.62) in Bern, Mittelland region, and the largest fold change estimate of 1.60 (95%CrI 1.11 to 2.33) in Schwyz, Central region. It was also spatially structured, with an estimated 36% of the variance attributable to the adjacency between WWTP catchment areas. Overall, the model was able to capture the dynamics of SARS-CoV-2 wastewater viral load in each WWTP with a coefficient of determination *R*^2^ of 0.60, with a few temporary local deviations from the regional time trends at the WWTP level (Supplementary Text S1).

Dynamic-time-warping distance-based clustering of these local deviations from the global temporal trend revealed notable geographical clustering in all phases except 4 (Figure 4A, Figure S19 (S1 Text)). The geographic signal in the communities became clearer when considering only the WWTPs whose median distance within cluster was lower than the lowest five percent of distances to WWTPs outside of the cluster (Figure S20 (S1 Text)).

**Fig 4.**
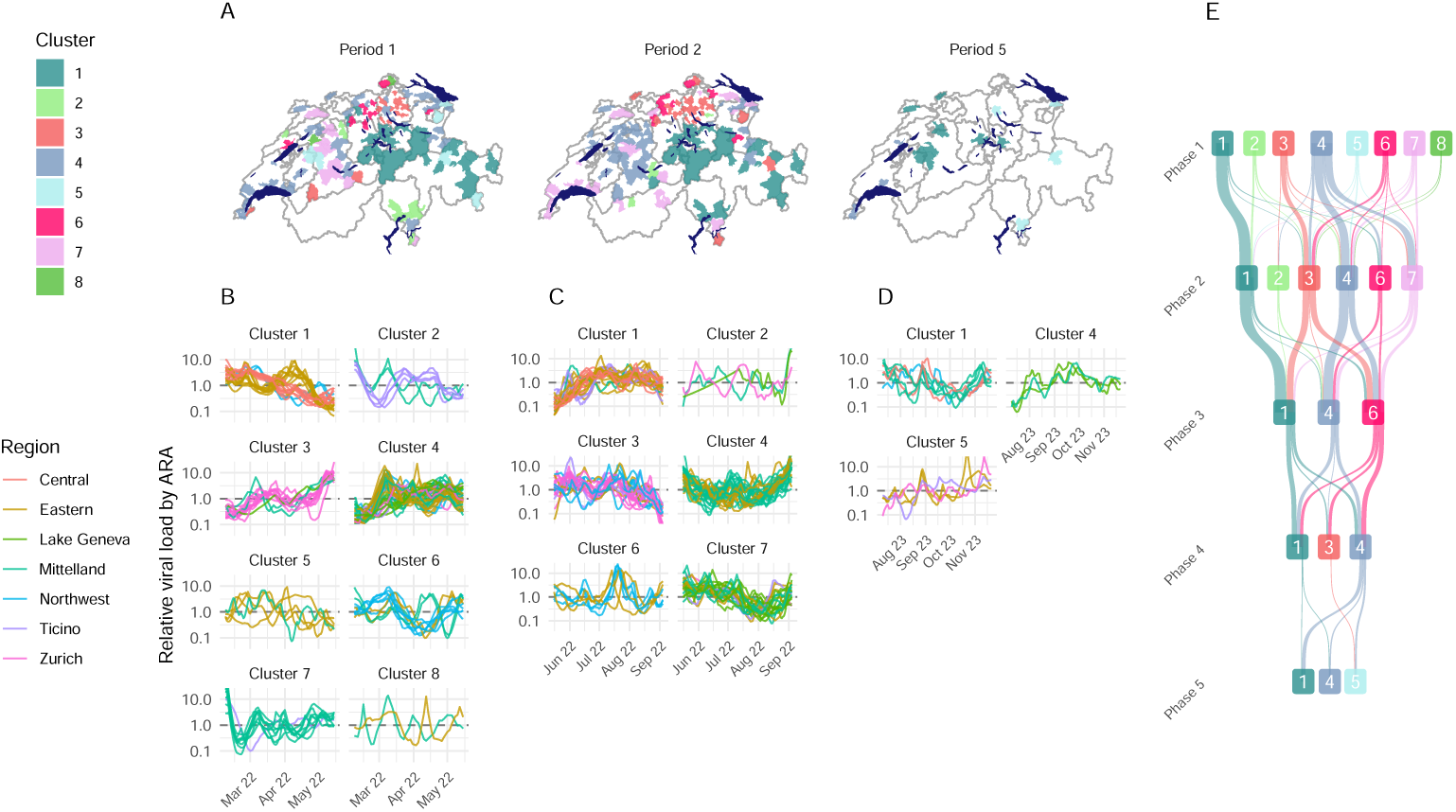
(A) Communities of WWTPs identified using hierarchical clustering of time-series in phases 1, 2 and 5 (B - D) time-series of local deviation from national wastewater concentration trends in their relevant clusters for phases 1, 2 and 5 respectively. E) Sankey plot describing how WWTPs flow between partitions in each analysis phase.

By cutting the dendrogram (Figure 5, S1 Text) at particular heights, we were able to investigate how communities form at different scales. In phases 1,2 and 3 the initial partition in two broadly divided eastern and western Switzerland. The region of Zurich was not consistently associated with either cluster. In phase 1, more urban areas tended to be combined with the cluster that is predominantly associated with Western Switzerland, which comprises most of the population. However, this pattern was less clear in phases 2 and 3.

**Fig 5.**
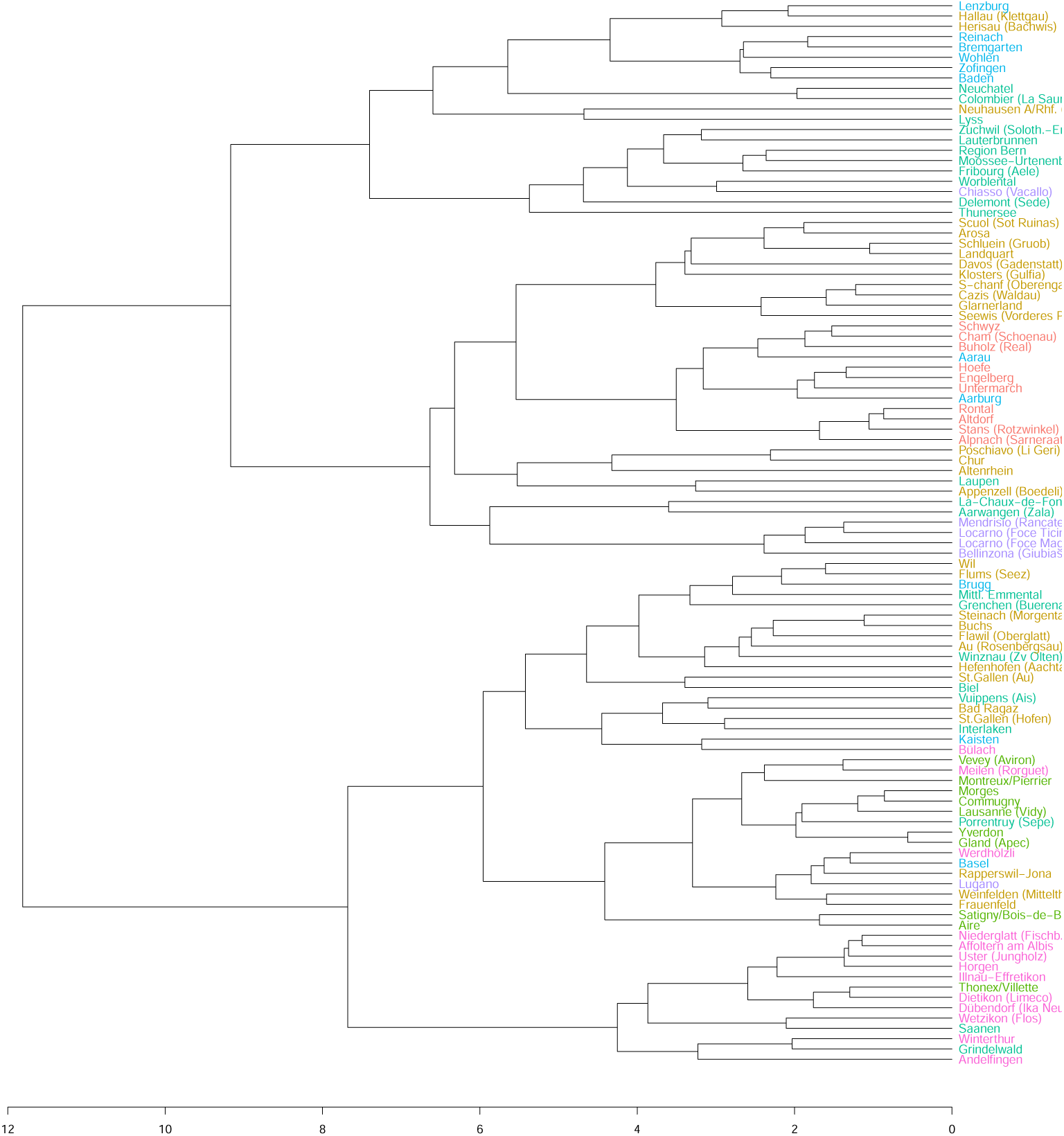
Unweighted pair group method with arithmetic mean (UPGMA) dendrogram based on dynamic time warping distance for Phase 1 of the analysis. Colour indicates region (NUTS2)

Considering higher order partitions, the communities in phase 1 remained strongly geographically clustered with as many as 8 communities defined. In particular strong geographic clusters were detected in the Central and Eastern regions (cluster 1), Zurich (cluster 3) and Lake Geneva region (cluster 4), all with relatively low normalised average pairwise distances (Table 2). Similar geographical structure remained in phases 2 and 3, however new clusters ceased to be geographically arranged when more than 4 or 5 partitions were defined. Notably, many of the strongest geographic clusters, for example Central and eastern regions (cluster 1), Zurich (cluster 3) and Lake Geneva region (cluster 7), remained present in phase 2. In addition, a strong geographic cluster (cluster 4) formed in the Mittelland region (Figure 4 C and E). Also notable during the earlier phases of monitoring was the relative proximity of the trajectories of the WWTPs that serve the three largest cities (Zurich, Geneva and Basel). In phase 1 all three cities were partitioned into the same cluster, along with Lugano (the largest city of the canton of Ticino) and the rest of the Lake Geneva region. In phase 2 Zurich and Basel were partitioned into a shared cluster and Geneva is partitioned with Lugano. By Phase 5 however, they were partitioned into three distinct clusters.

**Table 2.**
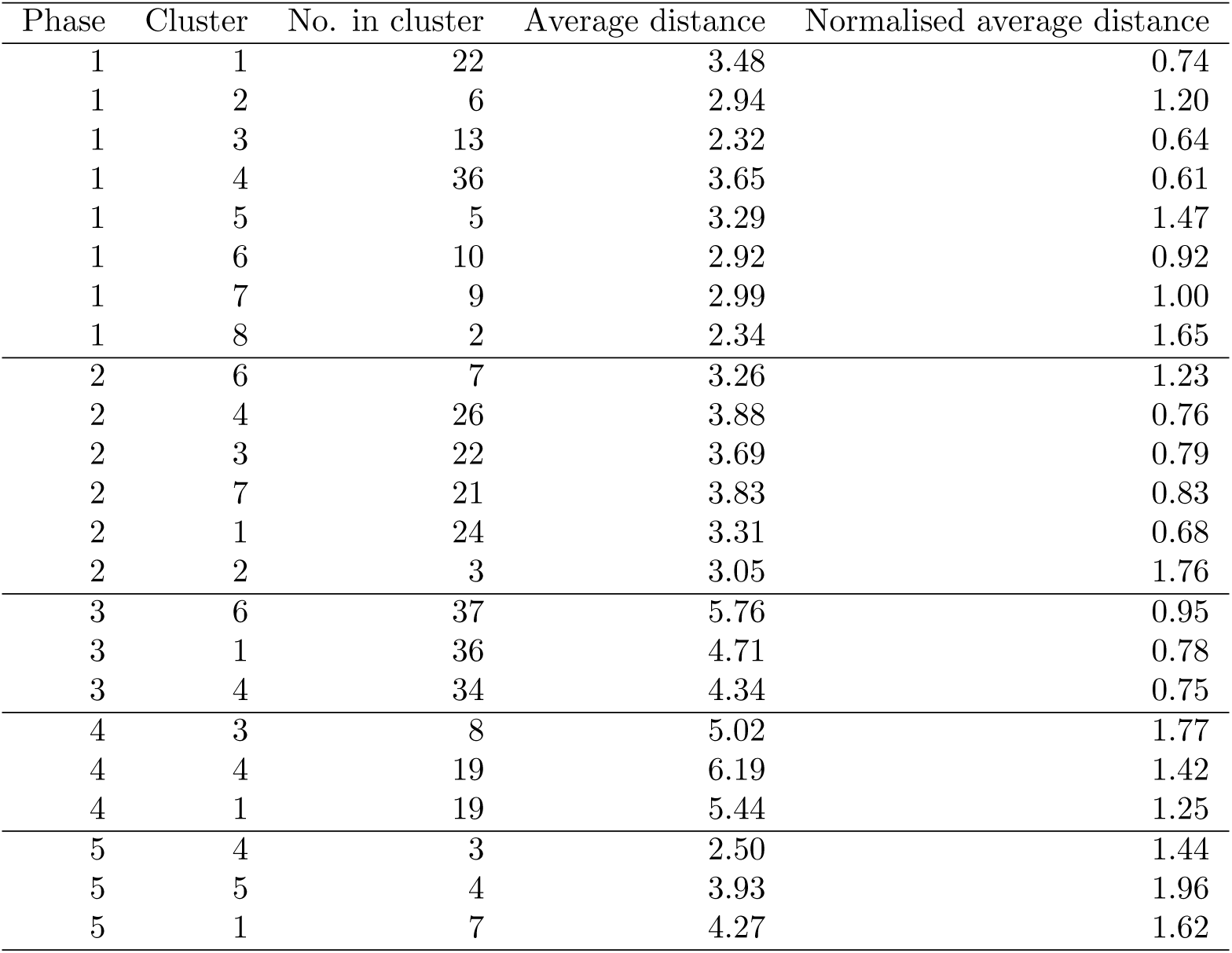
Size of and average distance between member WWTPs of clusters in each phases of the analysis.

| Phase | Cluster | No. in cluster | Average distance | Normalised average distance |
| --- | --- | --- | --- | --- |
| 1 | 1 | 22 | 3.48 | 0.74 |
| 1 | 2 | 6 | 2.94 | 1.20 |
| 1 | 3 | 13 | 2.32 | 0.64 |
| 1 | 4 | 36 | 3.65 | 0.61 |
| 1 | 5 | 5 | 3.29 | 1.47 |
| 1 | 6 | 10 | 2.92 | 0.92 |
| 1 | 7 | 9 | 2.99 | 1.00 |
| 1 | 8 | 2 | 2.34 | 1.65 |
| 2 | 6 | 7 | 3.26 | 1.23 |
| 2 | 4 | 26 | 3.88 | 0.76 |
| 2 | 3 | 22 | 3.69 | 0.79 |
| 2 | 7 | 21 | 3.83 | 0.83 |
| 2 | 1 | 24 | 3.31 | 0.68 |
| 2 | 2 | 3 | 3.05 | 1.76 |
| 3 | 6 | 37 | 5.76 | 0.95 |
| 3 | 1 | 36 | 4.71 | 0.78 |
| 3 | 4 | 34 | 4.34 | 0.75 |
| 4 | 3 | 8 | 5.02 | 1.77 |
| 4 | 4 | 19 | 6.19 | 1.42 |
| 4 | 1 | 19 | 5.44 | 1.25 |
| 5 | 4 | 3 | 2.50 | 1.44 |
| 5 | 5 | 4 | 3.93 | 1.96 |
| 5 | 1 | 7 | 4.27 | 1.62 |

All geographical structure appeared to be absent from the communities detected during phase 4. This indicates relatively homogenous behaviour across the country. However the geographic signal returned in phase 5 despite the small number of WWTPs monitored. Three clear communities existed: Zurich, the Eastern region and Ticino (cluster 5); Bern, Solothurn, Basel and the Central region (cluster 4) and around lake Geneva and Neucĥatel (cluster 5). Notably, clusters broadly organised by geographic regions that overlap with language differences. Cluster 1 speaking German with the exception of Porrentruy (French), cluster 5 speaking German with the exception of Lugano (Italian) and cluster 4 consisting of only French-speaking cantons.

## 4 Discussion

Our detailed analysis of population-normalised wastewater loads of SARS-CoV-2 RNA in Switzerland led to four important findings. First, we corroborated previous findings that viral load measurement can vary widely depending on laboratory methods [25], hampering the comparability of measurements done in different laboratories in the absence of methods aimed at adjusting for these biases. Second, the age structure of the population living in WWTP catchment areas influenced viral load measurements, as did to a lower extent the socio-economic level and the number of jobs. Third, the remaining variability in viral load across WWTPs was spatially structured, with the adjacency network between WWTPs explaining about a third of this variability. Fourth, statistical models with fixed effects and space-time components can be used to aggregate WWTP-level measurements and adjust for known and unknown but spatially structured sources of variability. In turn, this allows the production of more precise time trends at the regional level, that are highly correlated with SARS-CoV-2 hospitalizations.

Besides laboratory methods, we found that wastewater SARS-CoV-2 viral load was influenced by characteristics of the population living in WWTP catchment areas. Exploring potential factors, we only found a clear association with population age structure. The comparatively higher viral loads in WWTPs with more people aged above 65 (mirrored by lower viral loads in WWTPs with more people aged below 25, although it was compatible with no association) has implications on the understanding and interpretation of wastewater-based data. This effect is however consistent with studies reporting longer and more frequent faecal shedding upon SARS-CoV-2 infection in older people [22, 23, 38], which may bias estimates as older residents may on average shed virus into the wastewater system more frequently and for longer time periods, elevating concentrations relative to the incidence of infection. In that sense, higher SARS-CoV-2 wastewater viral loads in WWTPs with older populations should not be interpreted as indicative of higher incidence and/or prevalence in these regions, but rather as an artifact of population-structured shedding patterns. In a practical sense, this artifact may have the unexpected benefit of allowing for a more precise monitoring of viral circulation in older populations that are generally more vulnerable to infection.

Once these fixed effects are adjusted for, the remaining variability at the WWTP-level was small, with a ratio of 3 between the lowest and highest average viral load compared to 71 in the raw data. One possible explanation for the remaining heterogeneity is that the population declaring main residence within the limits of the WWTP catchment area may not entirely overlap with the population potentially engaged in wastewater faecal shedding for a variety of reasons, including commuting, tourism and other types of travelling, in every case going both directions. We attempted to measure this effect using the number of full-time equivalent jobs per population in each WWTP (the employment factor), but our results were not conclusive. The absence of a weekend effect also speaks against the influence of commuting. A closer look at the residual temporary deviations from the regional time trends at the WWTP level suggests tourism as a potential driver, with for instance higher viral loads in the highly touristic regions of Lauterbrunnen and Grindelwald during the summer of 2022 (Supplementary Text S1, Figure S13), however we were unable to formally account for population fluctuations as a result of tourism in this analysis. Future research may consider other types of indicators (e.g. hotel nights, public transport usage) to better understand the influence of population movements on WBS.

By quantitatively comparing the temporal structure of local deviations in viral load time-series, we were able to identify clear geographic clusters over much of the monitored period. In particular, the earliest phase showed the highest resolution of spatial clustering in viral load signals. The clustering continued to be present in later phases with much of the structure being preserved between phases, however the regions of similarity increased in size, both in terms of number of WWTPs and geographical region covered in later phases. This pattern suggests that transmission patterns may have become more mixed in later phases. This is consistent with relaxation of restrictions on movements as well as analyses of how contact patterns slowly returned to pre-pandemic levels in other parts of Europe [39]. The retention of geographic structure indicates however that key spatial transmission structures remain even in the post pandemic era, with the clusters in phase 5 largely dividing the population by language.

We identified several limitations in our study, which made use of data from a large national monitoring framework. For logistical reasons multiple laboratories were contracted to quantify viral concentrations concurrently. These laboratories were responsible for developing and implementing their own protocols. All laboratories with the exception of Eawag used qPCR which is reliant on a standard-curve to translate cycle thresholds to concentrations. The calibration of the standard curve can change as the variant mix in circulation varies over time. Changes in methodology were made to account for this variation in concentration quantification, which we account for by including the joint lab-method parameter. However, the accuracy of the calibration is likely to have varied over the course of their use, which would then be expected to directly impact the concentrations measured. Moreover, information provided by the various laboratories to the FOPH regarding changes in methods over time may be incomplete and completeness may vary between laboratories.

In our statistical analysis, we accounted for a number of effects to evaluate associations between various factors and measured viral concentrations. The model we have implemented allows for concentrations to change over time and space independent of these factors, however it does not allow for interaction between the factors and the spatio-temporal component of the model. This means that if the effects of the factors were to vary over the course of the study period or vary geographically, we would not capture these variation in our model. We contend that this is a reasonable simplification of the model in the context of the study setting and period. By nature, our study is also subject to ecological bias. This could affect in particular the correlation between viral load and COVID-19 hospitalizations.

Our findings have practical implications for future surveillance efforts involving SARS-CoV-2 and other respiratory viruses with faecal shedding. Whenever possible, WBS should centralize the laboratory analyses to a single laboratory, or improve the standardization of laboratory procedures. Alternatively, statistical models such as ours can be used to adjust for systematic differences between laboratories. We argue that this statistical processing of WWTP-level viral load measurement should be implemented in routine, as it allows to produce cleaner, more interpretable time trends that may be helpful to public health decision-makers, who are challenged by the high variability of raw viral load measurements processed in multiple laboratories.

## Data Availability

Data on viral concentration of SARS-CoV-2 in wastewater and data on COVID-19 hospitalizations are freely available from https://www.idd.bag.admin.ch/. Code is available on https://github.com/foph-modelling/multilevel_wbe.

https://github.com/foph-modelling/multilevel_wbe

## Supporting information

**S1 Text. Additional methods and results regarding the analysis of the sources of variation in viral load and the clustering of wastewater treatment plants based on time trends.**

## Acknowledgments

We wish to gratefully acknowledge the numerous personnel involved in the collection of wastewater samples at the many wastewater and processing in the following laboratories (Eawag, Amt für Lebensmittelsicherheit und Tiergesundheit Kanton Graubünden, Eurofins Scientific AG, Kantonales Labor Basel-Stadt, Kantonales Labor Zürich, Laboratorium der Urkantone, Microsynth AG, Service de la Consommation et des Affaires Vétérinaires du Canton de Neucĥatel, and University of Applied Sciences and Arts of Southern Switzerland).

