## Supplementary materials S1 Text for "Determinants and spatio-temporal structure of variability in wastewater SARS-CoV-2 viral load measurements in Switzerland: key insights for future surveillance efforts"

### S1.1 Analysis of sources of variation in viral load

#### S1.1.1 Additional statistical methods

Spatial aspects were added incrementally in the model of SARS-CoV-2 viral load (see section S1.1.2 below), leading to the selection of model 4 for the main results. Model 4 assumes that the viral load $y$ in WWTP $i$, region $j$ and day $t$ follows a Gamma distribution with:

$$y_{i,j,t} \sim \text{Gamma}(\exp(\mu_{i,j,t}), \eta), \tag{1}$$

where $\eta$ is a shape parameter and $\exp(\mu_{i,j,t})$ is the mean expected viral load. This mean is modelled using a linear predictor:

$$\mu_{i,j,t} = \alpha + \beta X + \omega_{j,t} + \zeta_i + \theta_{i,t} + \delta_{i,t} + \kappa_{i,t}, \tag{2}$$

where $\alpha$ corresponds to a unique intercept, and $\beta$ is a vector of slope parameters applied to the covariate matrix $X$ which included covariates for weekends (binary), national holidays (binary), laboratory method (11 levels, reference EAWAG), proportion of the population aged under 20 (continuous) or over 65 years old (continuous), employment factor (continuous) and median Swiss-SEP (continuous). All continuous covariates were centred and scaled, so $\exp(\beta)$ can be interpreted as relative increase in viral load for a change of 1 standard deviation in the covariate. Both $\alpha$ and $\beta$ were given weakly-informative normal priors with mean 0 and variance 5.

The model also included a random walk of order 2 to model the time trends of viral load over time in each of the seven regions:

$$\Delta^2\omega_{j,t} = \omega_{j,t} - 2\omega_{j,t-1} + \omega_{j,t-2} \sim \mathcal{N}(0, \tau_\omega^{-1}), \tag{3}$$

independently for each region $j$. The precision $\tau_\omega$ was given a log-gamma penalized complexity prior with shape 1 and rate 0.01.

At the lower level, the regional time trend was shifted up or down in each WWTP using a re-parameterised Besag-York-Mollié model (BYM), here represented by parameter $\zeta_i$, a spatially structured component that combines both structured and unstructured random effects:

$$\zeta_i = \sqrt{\frac{\phi}{\tau_\zeta}} u_i + \sqrt{\frac{1-\phi}{\tau_\zeta}} v_i \tag{4}$$

where:

- $u_i$ represents the spatially-structured effect, modeled using a conditional autoregressive prior;
- $v_i$ is the unstructured random effect (*i.i.d.*);
- $\tau_\zeta$ is an overall precision parameter;
- $\phi \in [0, 1]$ is a mixing parameter that controls the proportion of variability explained by the structured versus unstructured components.

More details about the structure of the BYM model can be found in [1]. We selected penalized complexity priors for both $\tau_\zeta$ and $\phi$, whose exact formulation depends on the neighboring structure. These priors imply that we do not expect effect sizes larger than 3 on the multiplicative scale within the spatial structure, and that we expect the proportion of variance explained by the spatially-structured component to be around 50%.

In addition to this WWTP-level shift, unique for the whole period, we used a random walk of order 1 to capture temporary deviations from the regional trend, so that:

$$\Delta\theta_{i,t} = \theta_{i,t} - \theta_{i,t-1} \sim \mathcal{N}(0, \tau_\theta^{-1}), \tag{5}$$

where the precision was also given a penalized complexity prior (log-gamma with shape 1 and rate 0,01).

Last, we included additional variability for measurements below the LOD and below the LOQ using *i.i.d.* random effects:

$$\delta_{i,t} \sim \mathcal{N}(0, \tau_\delta^{-1}) \tag{6}$$

and

$$\kappa_{i,t} \sim \mathcal{N}(0, \tau_\kappa^{-1}), \tag{7}$$

to account for the fact that measurements below the LOD or LOQ had increased variability.

The corresponding `R-INLA` code is:

```
INLA::inla(vl ~ 1 +
                                         # beta
                     f(weekend,model="linear",mean.linear=0,prec.linear=.2) +
                     f(hol,model="linear",mean.linear=0,prec.linear=.2) +
                     f(prop_under_20b,model="linear",mean.linear=0,prec.linear=.2)
                         +
                     f(prop_over_65b,model="linear",mean.linear=0,prec.linear=.2) +
                     f(ssep3_medb,model="linear",mean.linear=0,prec.linear=.2)  +
                     f(employment_factorb,model="linear",mean.linear=0,prec.linear
                         =.2) +
                     f(lab_method,model="linear",mean.linear=0,prec.linear=.2) +
                                         # omega
                     f(day,model="rw2", scale.model=TRUE, constr=TRUE,
                          group=NUTS2, control.group=list(model="iid"),
                          hyper=list(prec = list(prior = "pc.prec", param = c(1,
                              0.01)))) +
                                         # zeta
                     f(ara1,model="bym2",
                          graph=path_graph,
                          scale.model = TRUE, constr = TRUE,
                          hyper = list(theta1 = list("PCprior", c(1, 0.01)),
                                       theta2 = list("PCprior", c(0.5, 0.5)))) +
                                         # theta
                     f(day1,model="rw1", scale.model=TRUE, constr=TRUE,
                          group=ara2, control.group=list(model="iid"),
                          hyper=list(prec = list(prior = "pc.prec", param = c(1,
                              0.01))))  +
                                         # delta
                     f(below_loq,model="iid") +
                                         # kappa
                     f(below_lod,model="iid"),
                   data = ww_all, family = "gamma")
```

#### S1.1.2 Iterative model building

Model building was done iteratively, creating and comparing versions based on domain knowledge, model fit and WAIC [2]. All models were based on Gamma regression, with only changes in the construction of the linear predictor (Eq. 2).

##### S1.1.2.1 Model 1. Unique national trend and *i.i.d* WWTP-level shifts

Model 1 is a slightly simpler version of the model detailed in section S1.1.1:

$$\mu_{i,j,t} = \alpha + \beta X + \omega_t + \zeta_i + \theta_{i,t} + \delta_{i,t} + \kappa_{i,t}, \tag{8}$$

Compared to equation 2, we assume a unique temporal trend for the entire country ($\omega_t$ instead of $\omega_{j,t}$, still based on a random walk of order 2). The shift from the national trend at the WWTP-level is assumed to be independent and identically distributed ($i.i.d$) across WWTPs ($\zeta_i \sim \mathcal{N}(0, \tau_\zeta^{-1})$).

This formulation makes the model highly flexible, and able to capture the time series of viral load in all WWTPs (Figure S1 - the posterior predictive checks are very similar for all models and are therefore not shown afterwards). The unique national trend $\omega_t$ reflects the average of the different time trends in viral load across WWTPs, accounting for the covariates (Figure S2). The exponentiate of $\omega_t$ can be interpreted as a relative viral load, centered around 1. The estimated shift from this unique trend estimated in each WWTP (measured by $\zeta_i$), again expontentiated to be interpreted in terms of relative viral load, is mapped in Figure S3. The residuals deviations is captured by $\theta_{i,t}$. The largest residuals can be used to identify specificities at the WWTP-level (Figure S4).

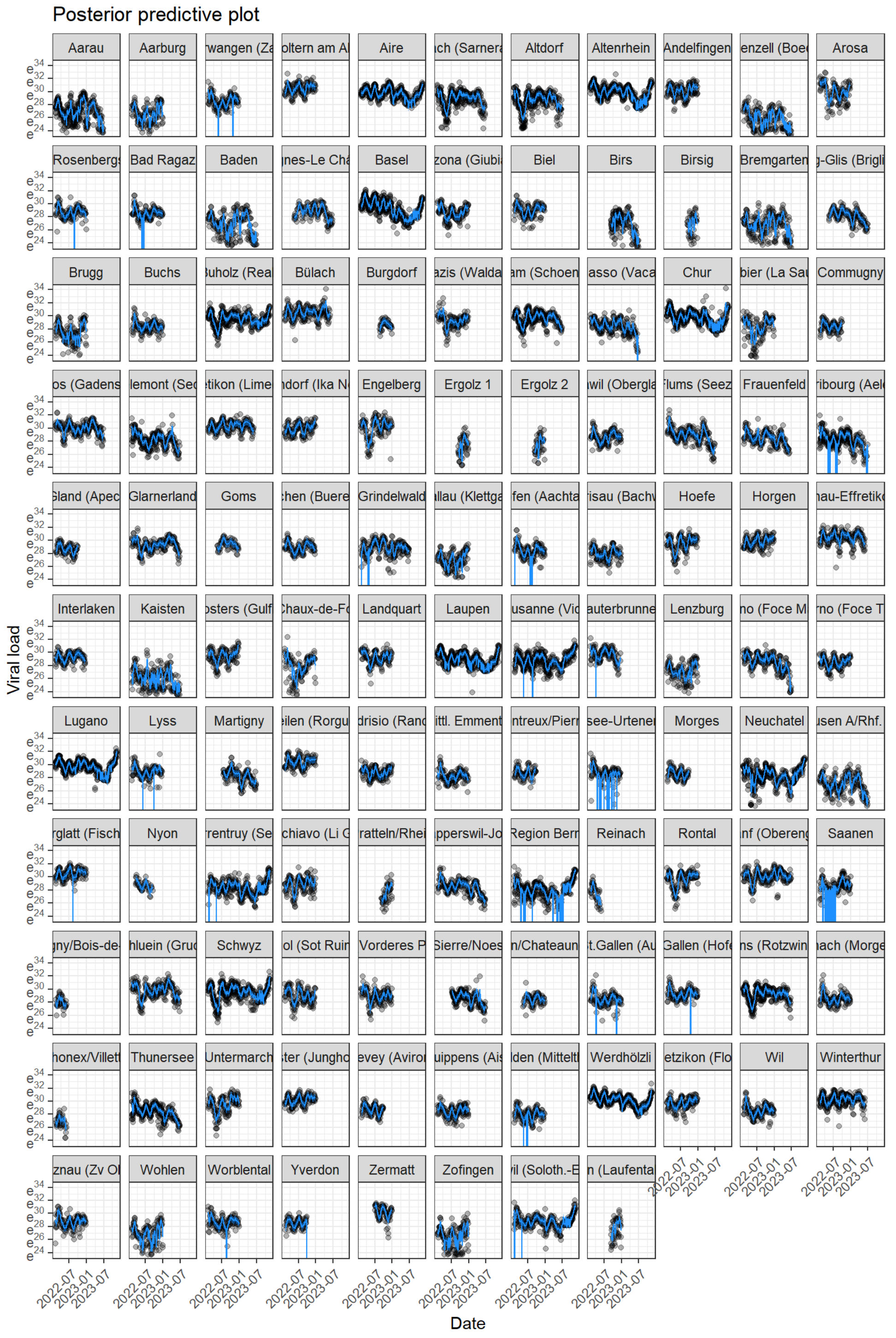

Figure S1: Posterior predictive plot for model 1.

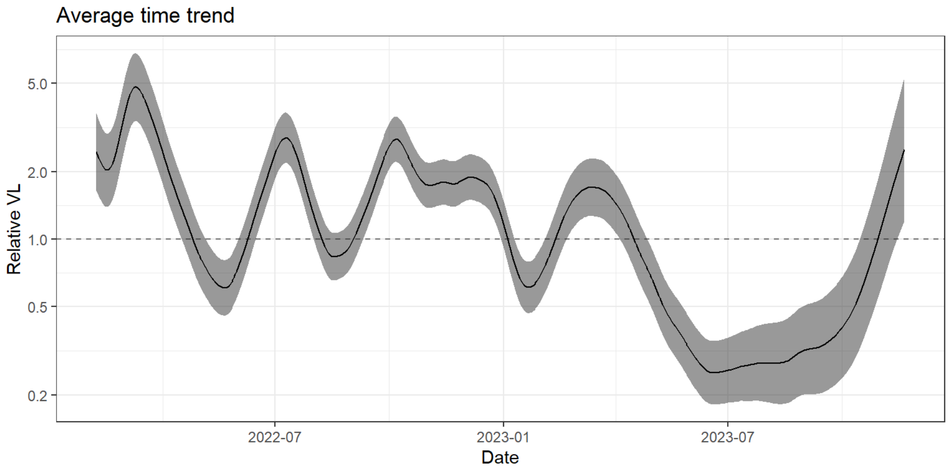

Figure S2: Posterior distribution of the estimated unique national trend for model 1 ($\exp(\omega_t)$).

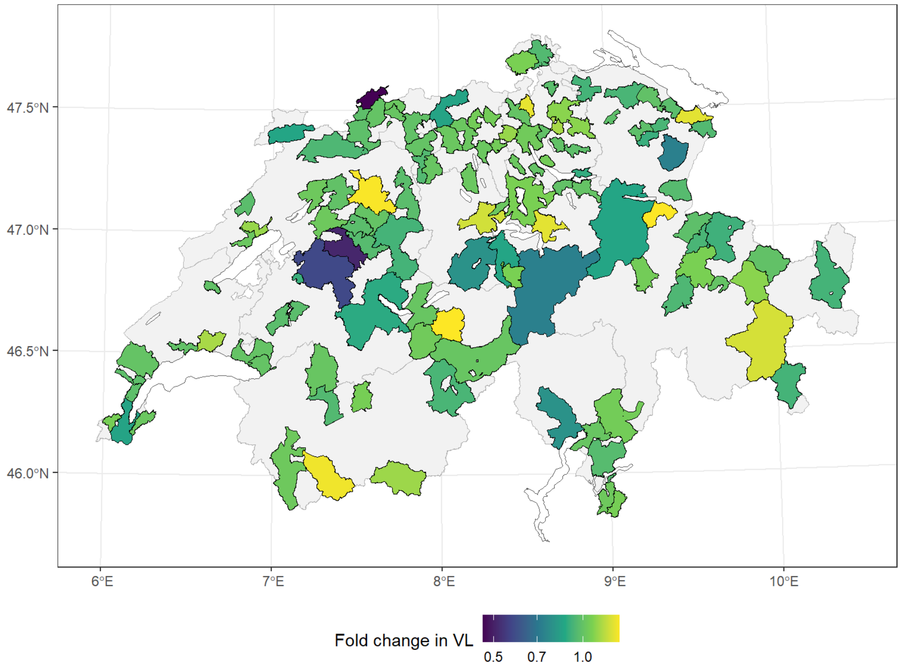

Figure S3: Posterior distribution of the estimated unique upward or downward shift from the higher-level trend in each wastewater treatment plant ($\exp(\zeta_i)$) for model 1.

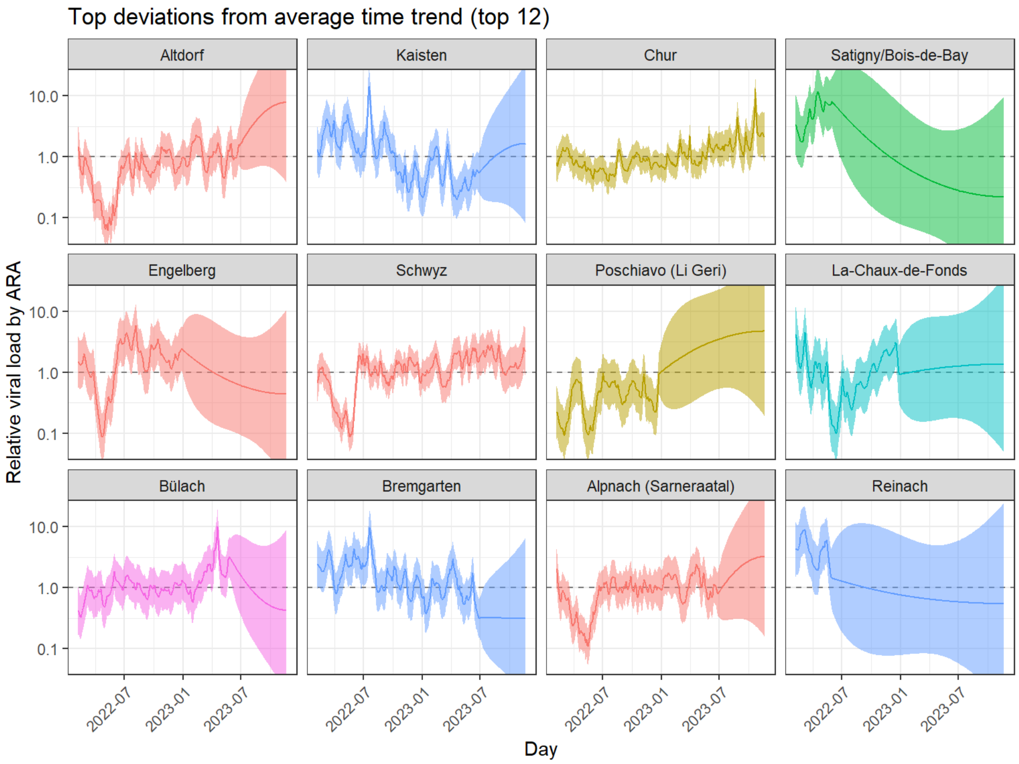

Figure S4: Posterior distribution of the top 12 residual temporary deviation from the higher-level trend in each wastewater treatment plant ($\exp(\theta_{i,t})$) for model 1.

##### S1.1.2.2 Model 2. Independent regional trends instead of unique national trend.

We now replace the unique national time trend with 7 independent time trends for each NUTS-2 region, as shown in equation 3. The overall shape of the average time trend is similar, but this allows for the identification of differences across NUTS-2 regions (Figure S5). The residuals at the WWTP-level are modified, as regional trends are able to capture part of the spatial variation (Figure S7). This change also improves the model fit as measured by the WAIC (Table S1).

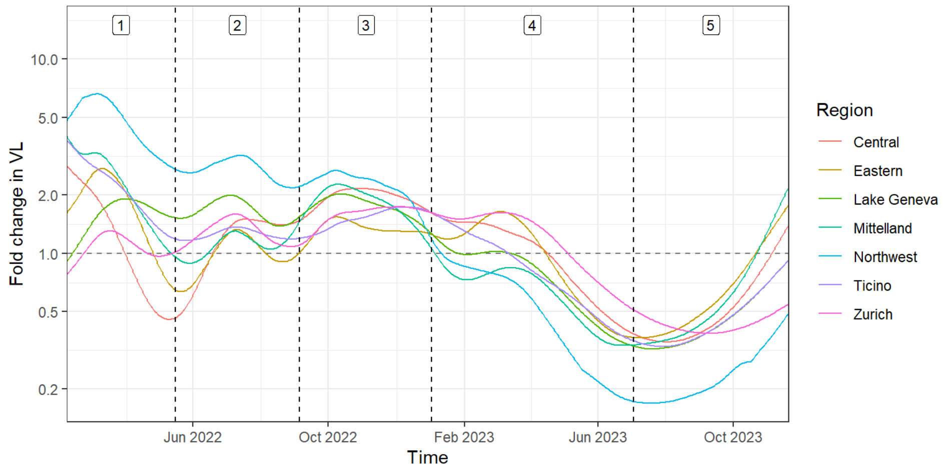

Figure S5: Posterior distribution of the estimated regional trends for model 2 ($\exp(\omega_{j,t})$).

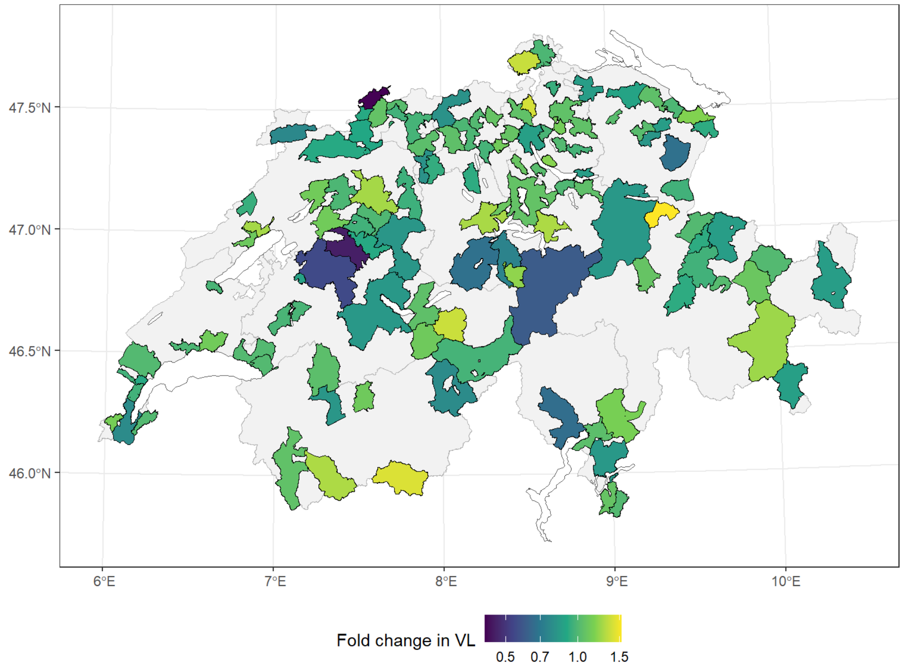

Figure S6: Posterior distribution of the estimated unique upward or downward shift from the higher-level trend in each wastewater treatment plant ($\exp(\zeta_i)$) for model 2.

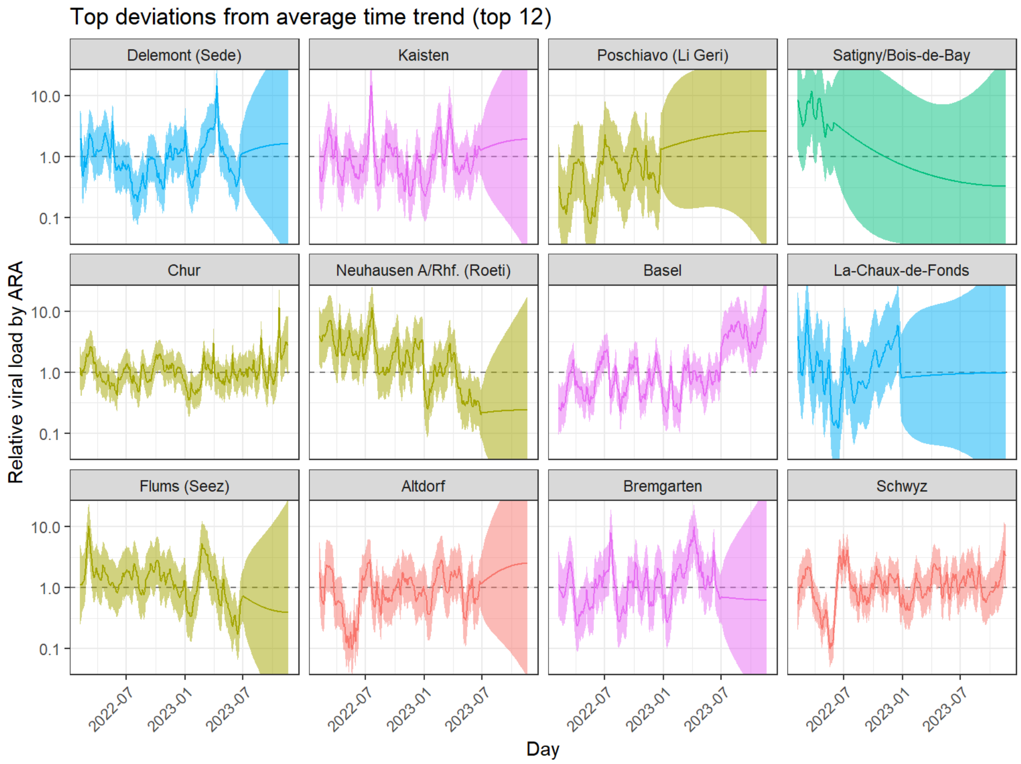

Figure S7: Posterior distribution of the top 12 residual temporary deviation from the higher-level trend in each wastewater treatment plant ($\exp(\theta_{i,t})$) for model 2.

##### S1.1.2.3 Model 3. Spatially structured WWTP-level shifts instead of *i.i.d*

Moving back to model 1 with its unique national trend, we now add a spatial structure on the WWTP-level shifts, as shown in equation 4. The national time trend is very similar to that of model 1 (Figure S8). This leads to more variability in the parameter estimates measuring WWTP-level shift (Figure S9) or residual deviations (Figure S10).

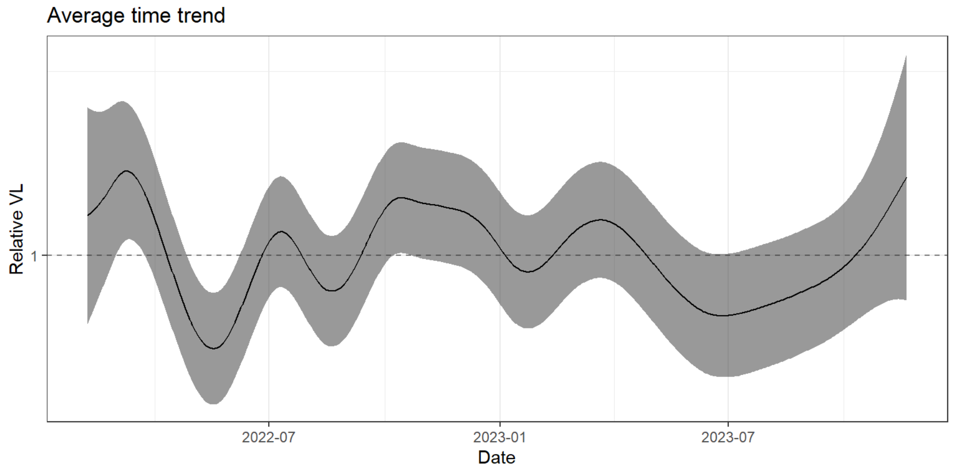

Figure S8: Posterior distribution of the estimated national trend for model 3 ($\exp(\omega_t)$).

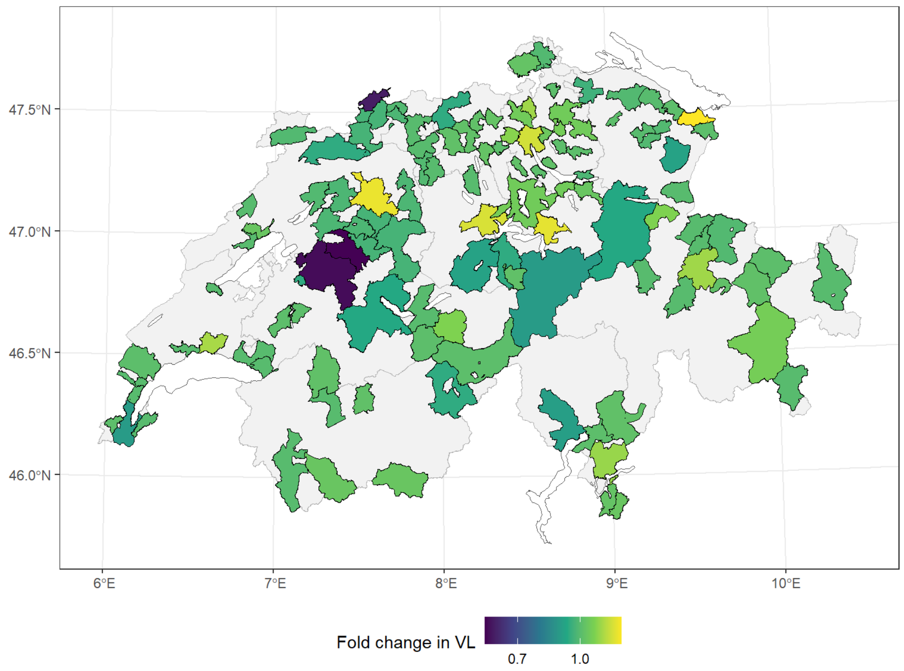

Figure S9: Posterior distribution of the estimated unique upward or downward shift from the higher-level trend in each wastewater treatment plant ($\exp(\zeta_i)$) for model 3.

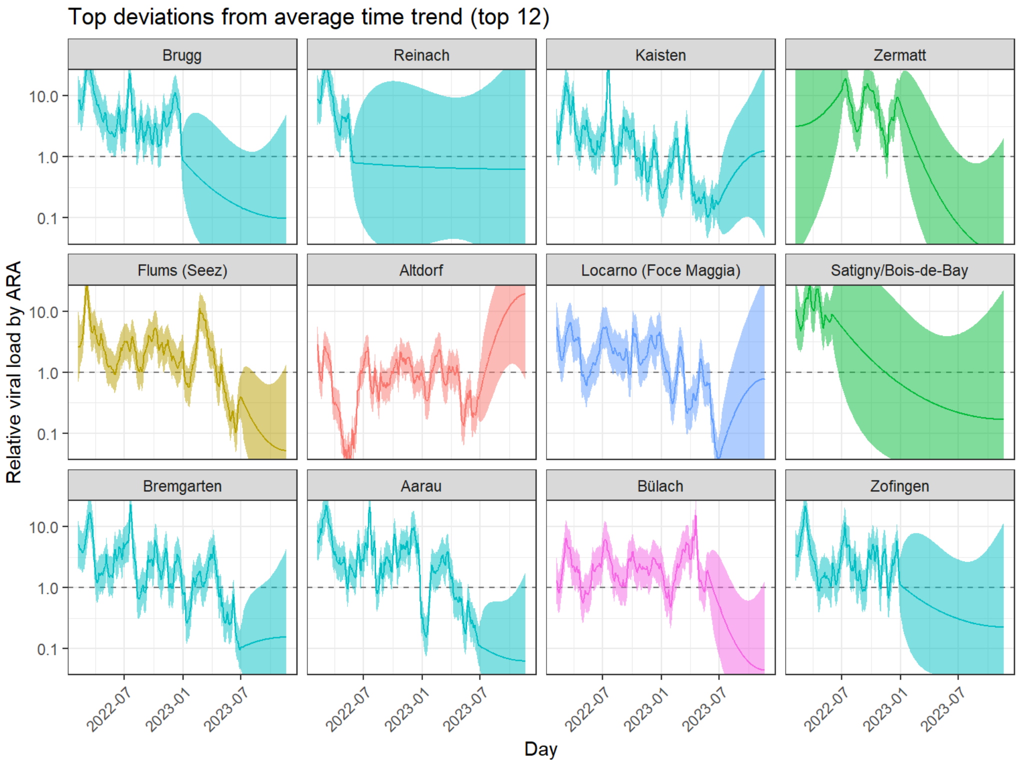

Figure S10: Posterior distribution of the top 12 residual temporary deviation from the higher-level trend in each wastewater treatment plant ($\exp(\theta_{i,t})$) for model 3.

##### S1.1.2.4 Model 4. Independent regional trends and spatially structured WWTP-level shifts

Model 4 combines the features added to model 2 and model 3, with independent regional trends and spatially structured WWTP-level shifts. The addition of the geographical neighborhood structure did not improve the model fit as measured by the WAIC, compared to model 2. However, we still retained this feature for the final model, as it allowed for a better smoothing of the WWTP-level shift. For the further explorations of the spatial structuring of SARS-CoV-2 viral load time series shown in section **??**, we used the outputs of model 1, as it did not include any spatial structuring.

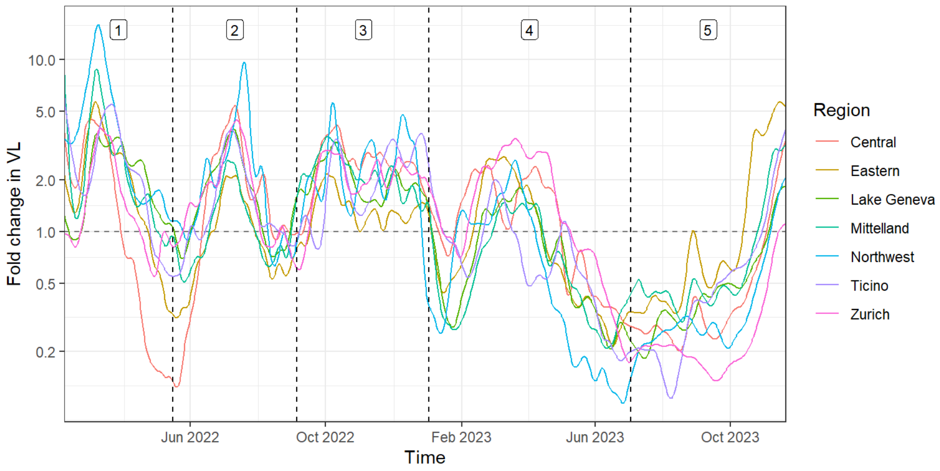

Figure S11: Posterior distribution of the estimated regional trends for model 4 ($\exp(\omega_t)$).

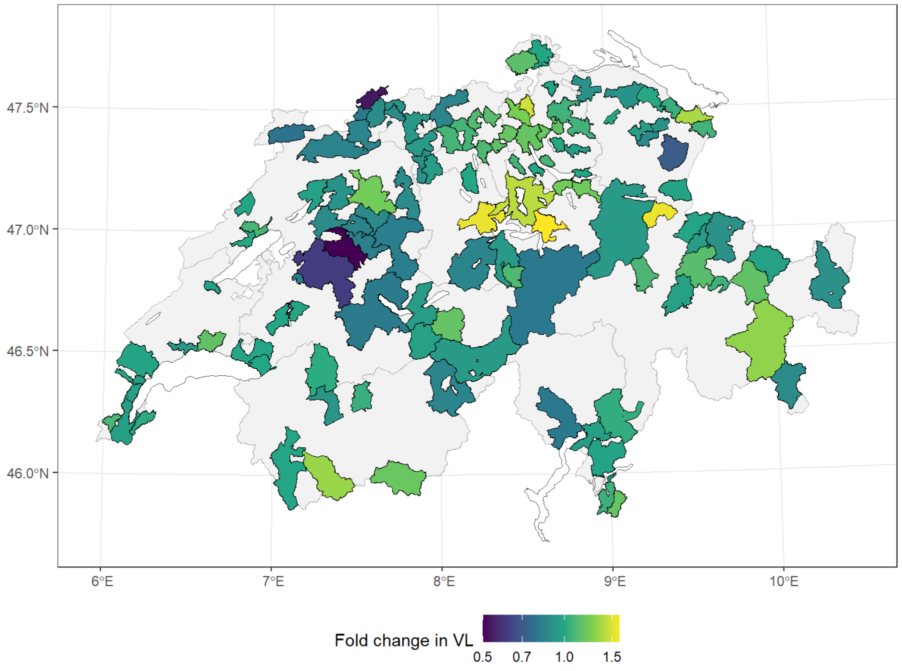

Figure S12: Posterior distribution of the estimated unique upward or downward shift from the higher-level trend in each wastewater treatment plant ($\exp(\zeta_i)$) for model 4.

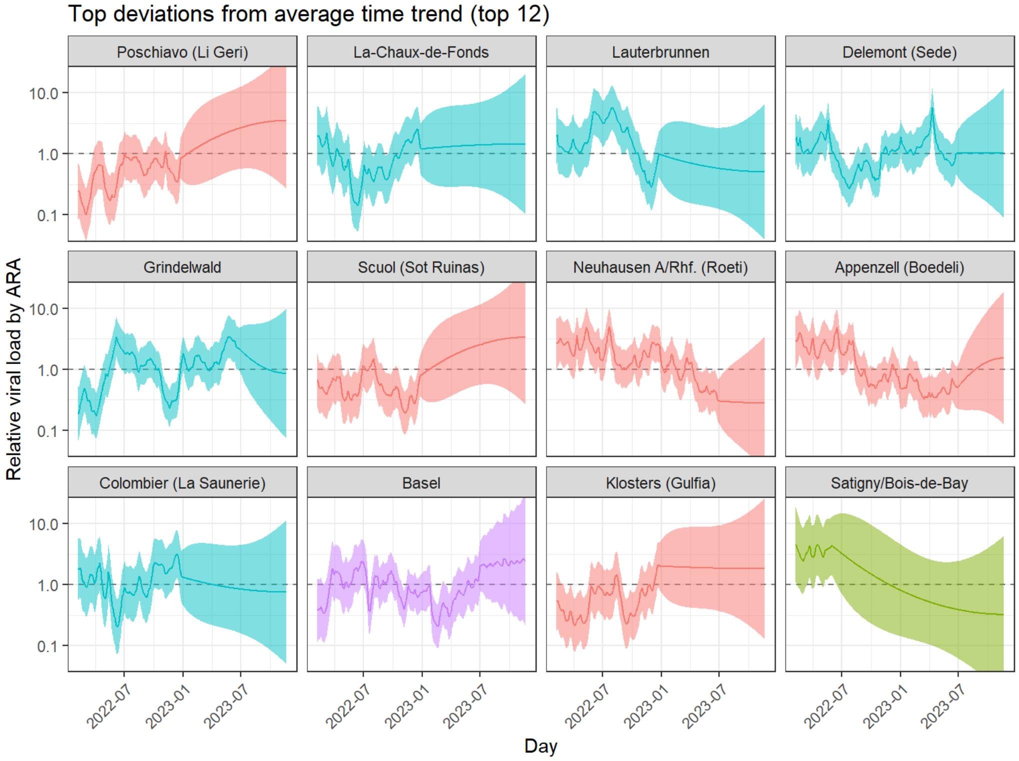

Figure S13: Posterior distribution of the top 12 residual temporary deviation from the higher-level trend in each wastewater treatment plant ($\exp(\theta_{i,t})$) for model 4.

Table S1: Summary of models and WAIC.

| Model | Description | WAIC |
|---|---|---|
| 1 | Unique national trend and *i.i.d.* WWTP-level shifts | 1,364,289 |
| 2 | Independent regional trends instead of unique national trend | 1,364,041 |
| 3 | Spatially structured WWTP-level shifts instead of *i.i.d* | 1,364,272 |
| 4 | Independent regional trends and spatially structured WWTP-level shifts | 1,364,061 |

#### S1.1.3 Correlation with hospitalisations

In order to assess the relevance of the time trends in viral loads by region obtained using model 4, we computed the Pearson correlation between $\omega_{j,t}$ and weekly counts of laboratory-confirmed SARS-CoV-2 hospitalisations in the same region, by period, obtained from the Federal Office of Public Health. We found a high degree of correlation overall (median 0.76). To quantify the added value of the model-based trends compared to the crude time series, we also computed the correlation between these and hospitalisations, by period. Correlation with crude time series were much lower (Figure S14B), which highlights the improvement obtained with the proposed model.

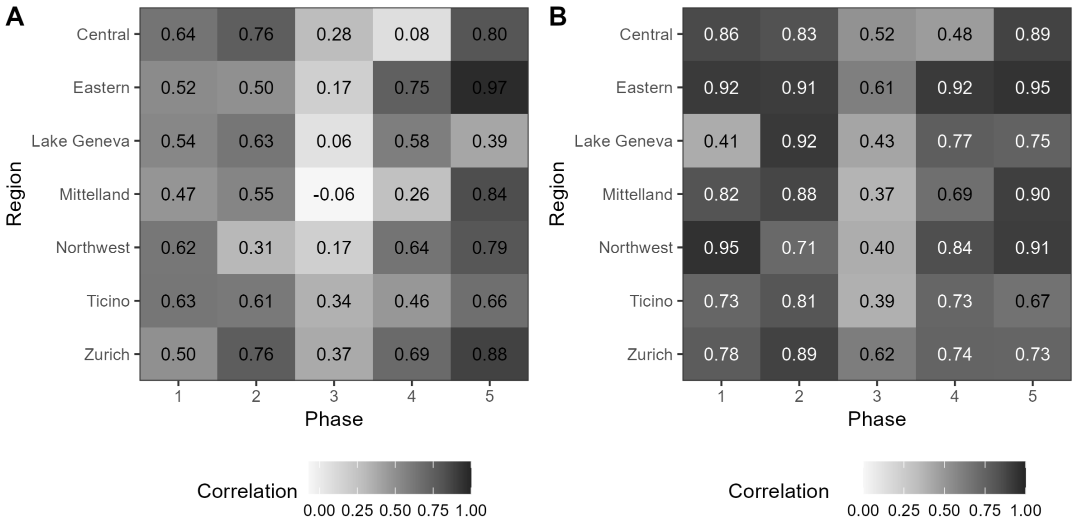

Figure S14: Correlation between unadjusted (panel A) or adjusted (panel B) time trends in viral load and laboratory-confirmed SARS-CoV-2 hospitalisations in the same NUTS-2 region.

In addition, we computed the cross-correlation between the adjusted time trends and laboratory-confirmed SARS-CoV-2 hospital admissions after shifting the time series of hospital admissions by a delay of -3 to +3 weeks (Figure S15). The results about the maximal correlation coefficients achieved for each region and each period indicate that the regional time trends were mostly synchronous (maximal delay of 0) or 1 week ahead (maximal delay of +7) of hospitalizations during periods 1, 2 and 3. During periods 4 and 5, as hospital admissions becomes less frequent, the level of synchronicity decreases.

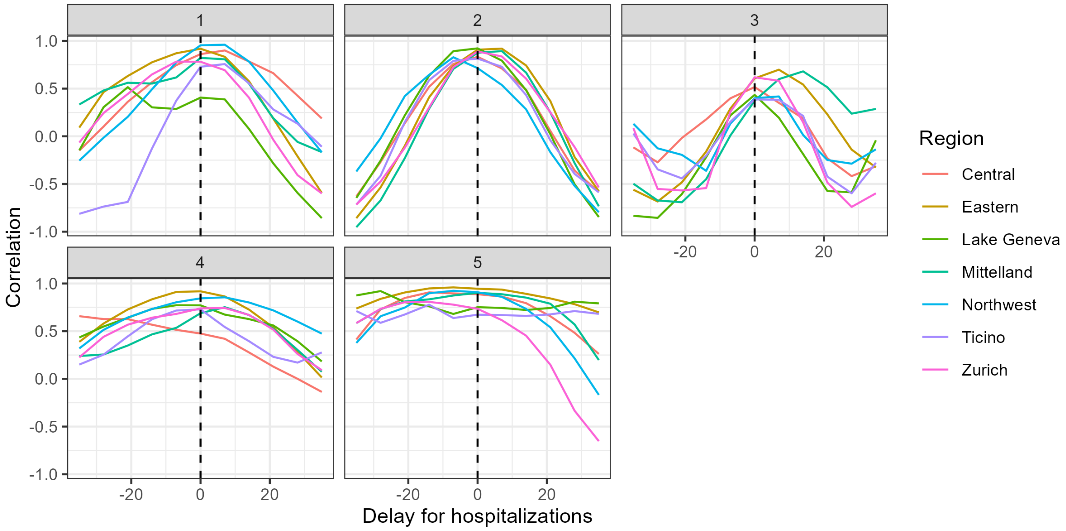

Figure S15: Correlation between adjusted time trends in viral load and laboratory-confirmed SARS-CoV-2 hospitalisations in the same NUTS-2 region shifted by -28 to +28 days (moving towards the right of the x-axis – a positive delay – means that the time trend of hospitalizations is shifted back in time).

### S1.2 Clustering of WWTPsb ased on adjusted temporal trends in viral loads

#### S1.2.1 Dendograms from the UPGMA clustering of each epidemic phase

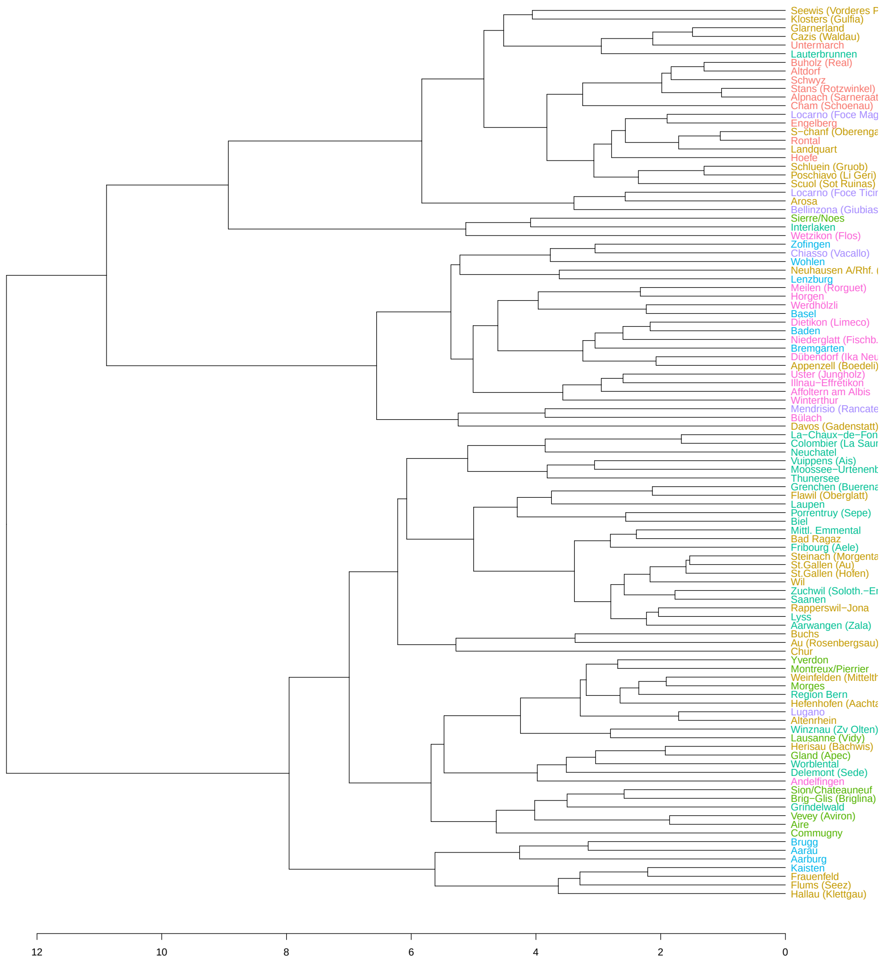

Figure S16: UPGMA dendrogram based on dynamic time warping distance for Phase 2 of the analysis

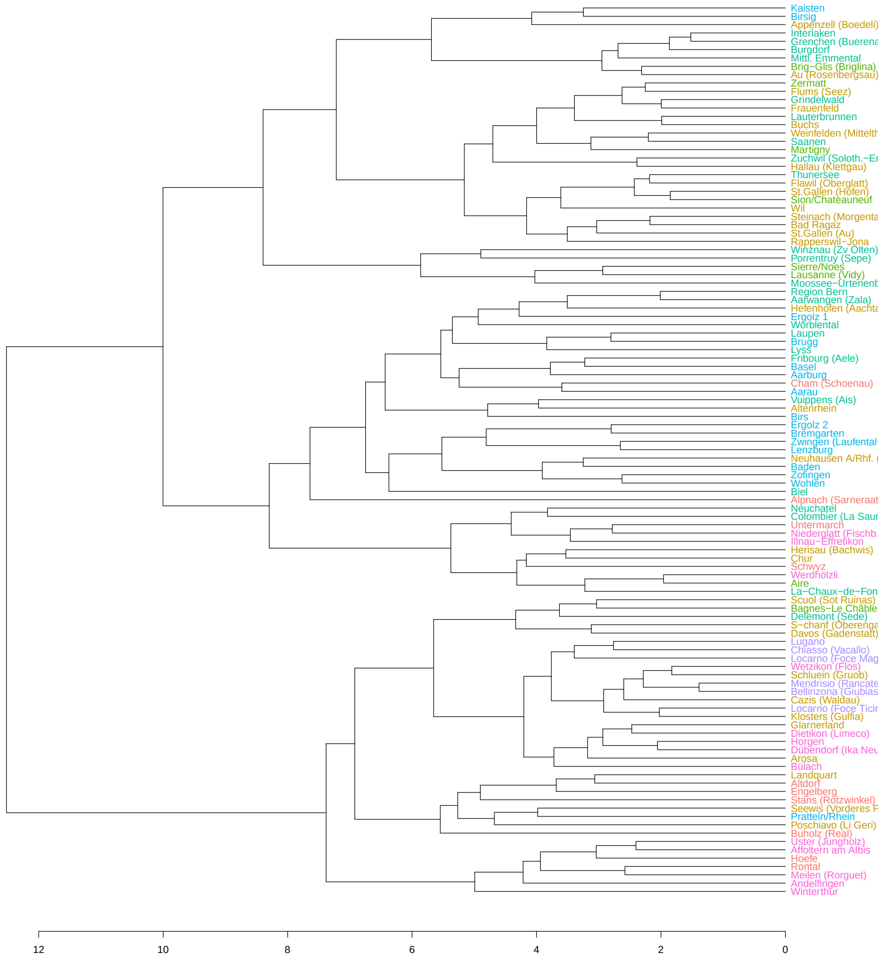

Figure S17: UPGMA dendrogram based on dynamic time warping distance for Phase 3 of the analysis

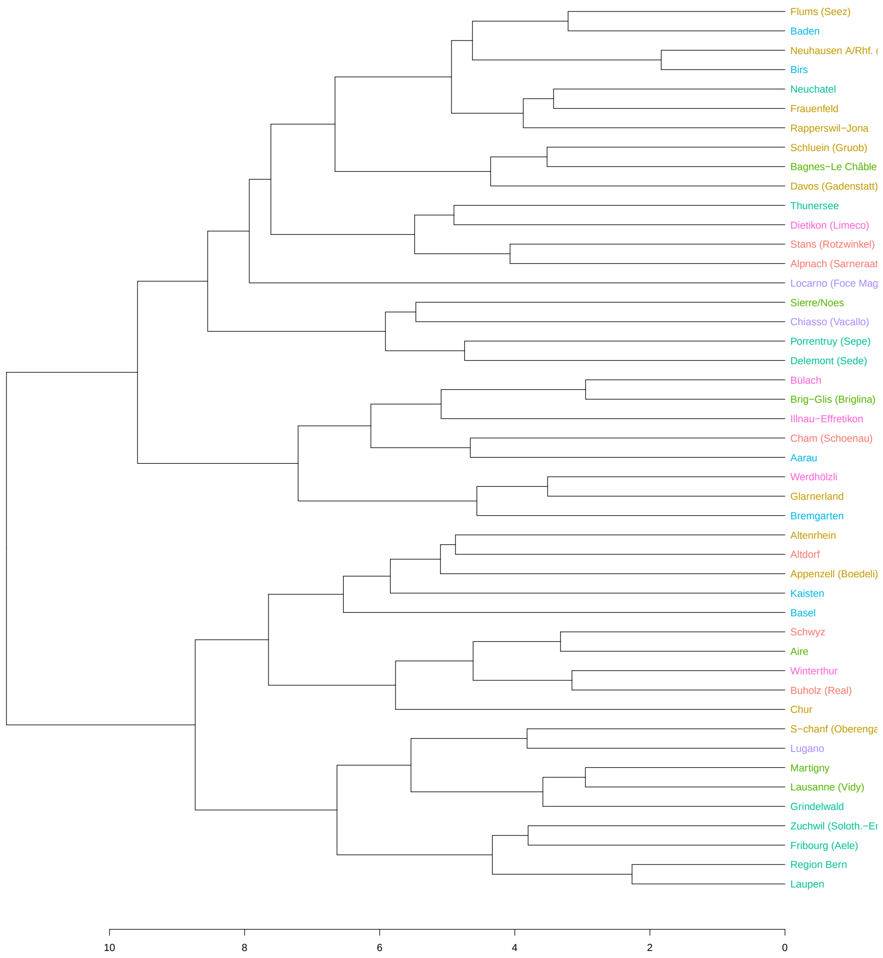

Figure S18: UPGMA dendrogram based on dynamic time warping distance for Phase 4 of the analysis

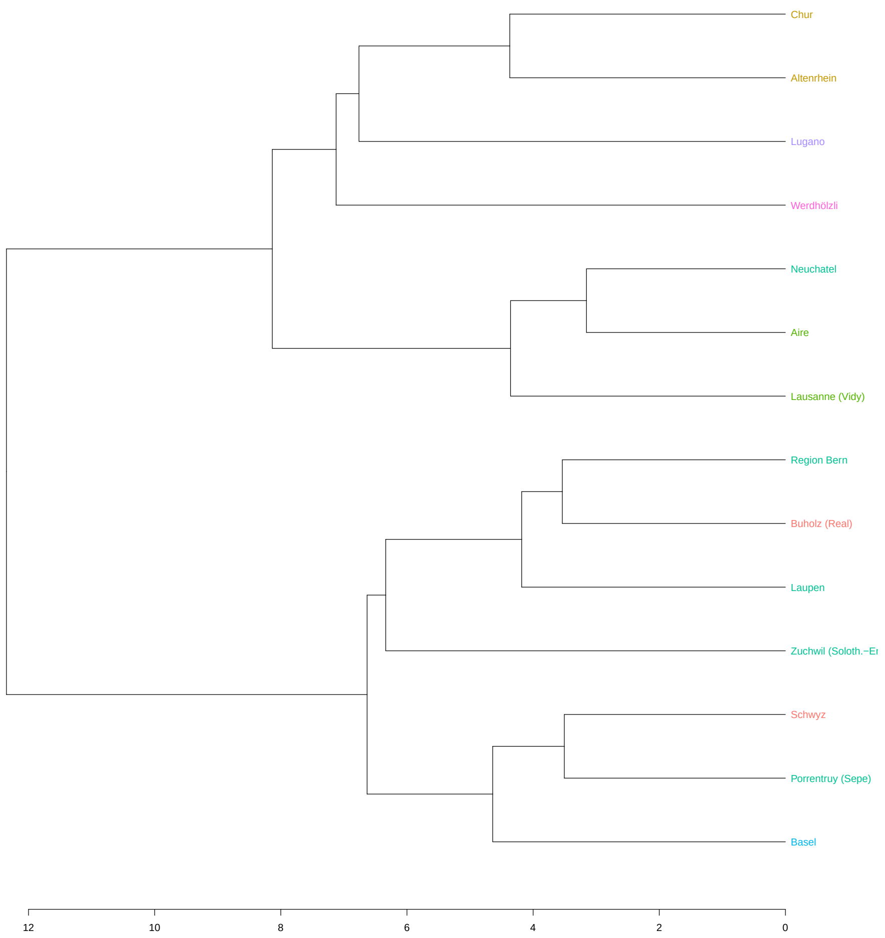

Figure S19: UPGMA dendrogram based on dynamic time warping distance for Phase 5 of the analysis

#### S1.2.2 Supplementary cluster maps

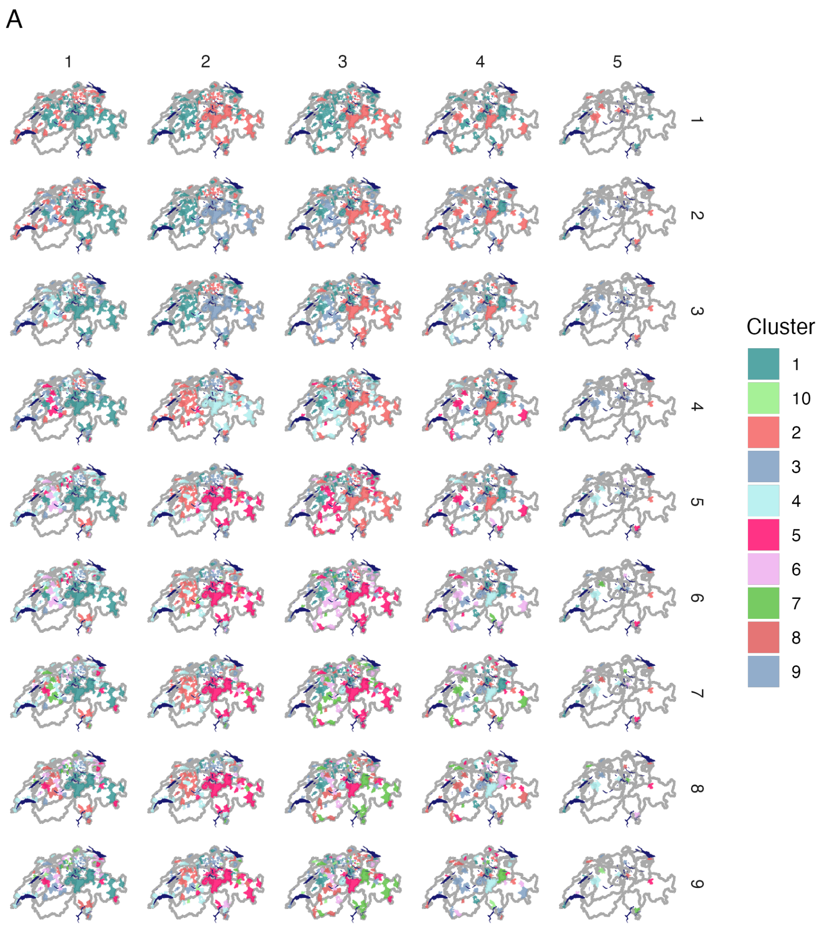

Figure S20: Clusters in each phase for k-clusters of 2 to 10

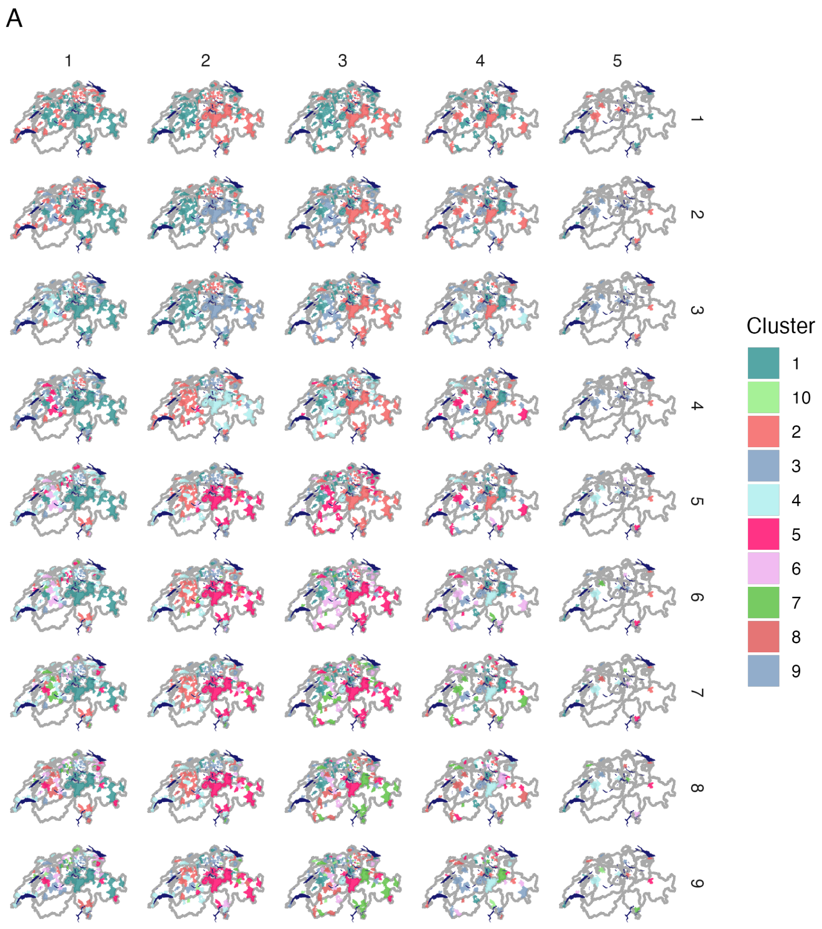

Figure S21: Clusters in each phase for k-clusters of 2 to 10 - showing only WWTPs strongly connected to their cluster

#### S1.2.3 Distance between clusters (centroids

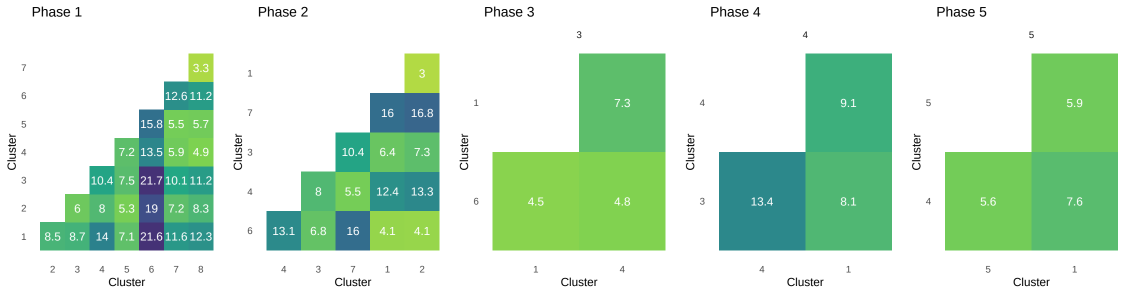

Figure S22: distance matrix for centroids of clusters in each phase

#### S1.2.4 All clusters series plots

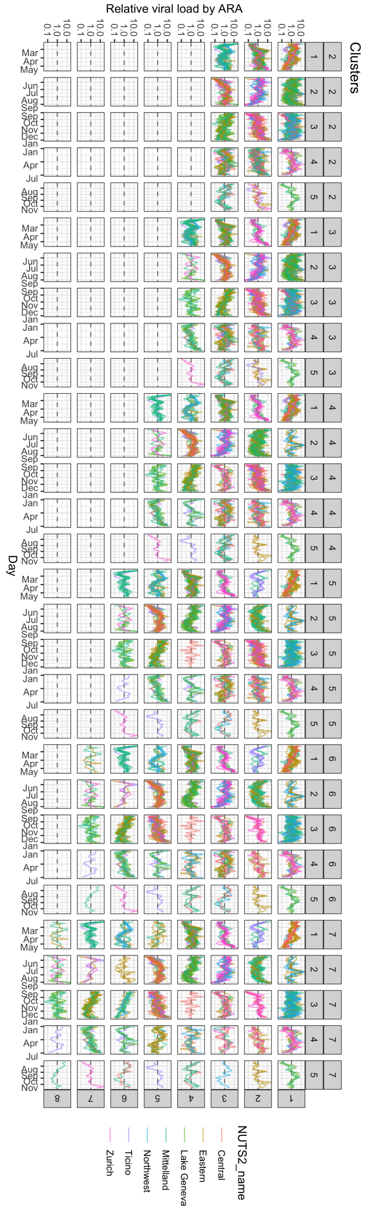

Figure S23: timeseries of each WWTP in each phase for k-clusters of 3 to 8
